# Aging affects ciliated cells development in the human endometrial epithelium

**DOI:** 10.1101/2023.05.22.23290333

**Authors:** Marina Loid, Darina Obukhova, Keiu Kask, Alvin Meltsov, Kasper Derks, Signe Altmäe, Merli Saare, Maire Peters, Ave Minajeva, Priit Adler, Kaarel Krjutškov, Masoud Zamani Esteki, Andres Salumets

**Affiliations:** Department of Obstetrics and Gynecology, Institute of Clinical Medicine, University of Tartu, Tartu, Estonia; Competence Centre on Health Technologies, Tartu, Estonia; Department of Clinical Genetics, Maastricht University Medical Center+, Maastricht, The Netherlands; Department of Biochemistry and Molecular Biology, University of Granada, Granada, Spain; Division of Obstetrics and Gynecology, Department of Clinical Science, Intervention and Technology, Karolinska Institutet and Karolinska University Hospital, Huddinge, Stockholm, Sweden; Institute of Biomedicine and Translational Medicine, University of Tartu, Tartu, Estonia; Institute of Computer Science, University of Tartu, Tartu, Estonia

## Abstract

The twenties are typically considered the prime reproductive years for women. However, in today’s modern world, many women are choosing to delay family planning, resulting in an increase of females in their forties seeking fertility treatment. Although *in vitro* fertilization (IVF) with donated oocytes and preimplantation genetic testing may help to address the impact of maternal age, the success rate for IVF treatment in this age group is still significantly lower. While endometrial changes, such as abnormal endometrial thickness, inflammatory background, and altered hormone response signaling, are associated with aging, little is known about the molecular features of endometrial aging and their impact on the ability to support embryo implantation. To better understand age-specific changes, we performed endometrial transcriptome profiling of young and advanced age females, undergoing hormonal replacement therapy (HRT) before frozen embryo transfer, followed by immunohistology analysis and single-cell-based deconvolution. Here, we identified 491 differentially expressed genes pointing to the effect of aging on decidualization, cell signaling, inflammation and endometrial receptivity. Our results indicate that p16^INK4a^ may be involved in cellular senescence and the suppression of metabolic and inflammatory processes essential for endometrial preparation for embryo implantation. We have also shown that the proportion of ciliated cells along with ciliary processes is affected by endometrial aging. These findings have important implications for future strategies aimed at improving infertility treatment in women of advanced reproductive age.

## Introduction

Nowadays, women often delay having children, which can have advantages due to their improved financial and social status. In 2021, mean maternal age in Europe was 31.1 years old (European Statistical System, 2022, full statistics can be seen at https://ec.europa.eu/eurostat/databrowser/view/tps00017/default/table?lang=en). The fertility rate decreases significantly in the late thirties due to a decline in ovarian reserve, which causes more women every year to seek medical assistance to get pregnant. Advanced assisted reproduction techniques (ART), such as *in vitro* fertilization (IVF) using oocyte donors have made procreation possible for women wishing to have children at almost any age^1^.

However, maternal age also negatively impacts IVF implantation rates when it comes to ART. The debate on whether endometrium ages has taken decades of research with still contradicting results. The main limitation arises from the prevailing knowledge of female general fertility decline at the age of 35, which refers to the progressive decline in ovarian function and subsequent hormonal changes. In endometrial tissue, epithelial cells accumulate mutations in cancer genes that increase with age^2^. Studies analysing women undergoing HRT have shown no significant difference in treatment outcomes between oocyte recipients younger and older than 39 years^3^. Although up to two-thirds of implantation failures can be avoided by preimplantation genetic testing for aneuploidies (PGT-A) of embryos prior to transfer, this rate drops significantly in women in their late forties, even when using donor embryos^4–6^. The reason may lie in significant molecular changes associated with tissue aging, not present in the receptive endometrium of younger women. This hypothesis is supported by the study showing higher pregnancy rates in ART cycles with younger gestational carriers compared to patients in both PGT and non-PGT groups^6^. Also, a recent retrospective study utilizing *in silico* analysis of previously collected endometrial gene expression data suggests the presence of molecular changes associated with age in women over 35 of years. However, the study analyzed only naturally cycling women, whose endometrial molecular profile is regulated by natural hormone dynamics^7^.

Previous studies have shown that not only gene expression can modulate the receptivity of endometrial tissue, but also the proportion of different cell types in the endometrium affects the embryo-endometrium interactions^8^. Distinguishing between cell types based on different molecular techniques can be challenging and the computational deconvolution method employed in our study is a steady approach to analyse endometrium as a dynamic pattern of cellular composition. In this study, we examine the molecular and cellular changes that occur in endometrial tissue in advanced maternal age (AMA) compared to young maternal age (YMA) groups and speculate on their significance for endometrial regeneration and receptivity formation. Also, the current study is the first one to utilize RNA sequencing to compare the transcriptomic profiles of endometrial samples from distant age groups.

## Results

### Genes and pathways significantly affected by AMA

To analyze the differences in gene expression between young and advanced maternal age endometria, total RNA samples from endometrial tissue of 12 YMA and 12 AMA women were sequenced. An overview of the study is presented in Fig.1. The main characteristics of study groups are presented in Fig.2a. In total, 19,357 mRNA transcripts were expressed in the analyzed endometrial samples. Based on gene expression profiles, the study samples clustered together with no clear outliers in both groups. The PCA based on gene expression data is shown in Fig.2b.

**Fig.1.**
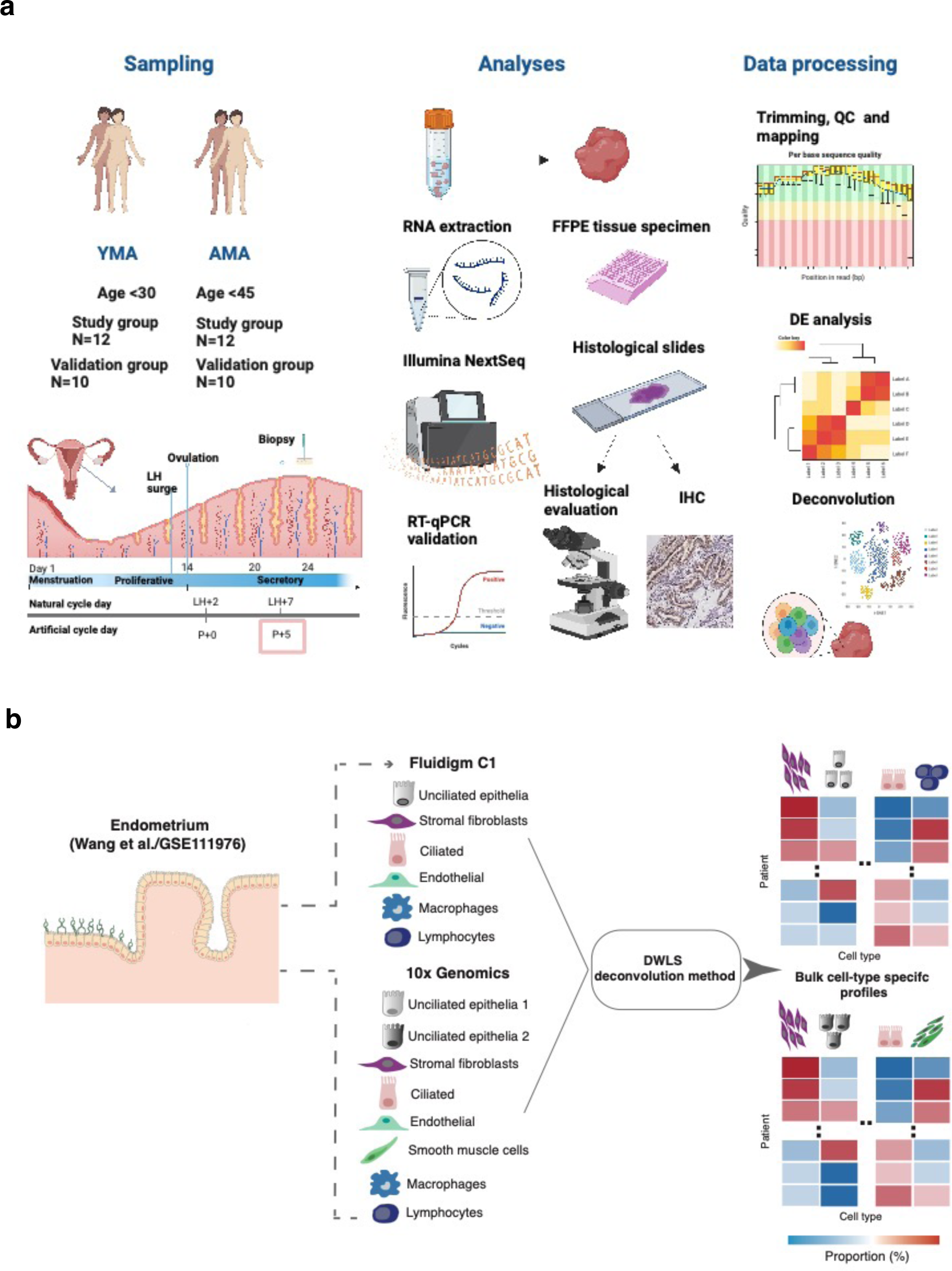
Schematic overview of the study design. **a**. The workflow of the analysis. Endometrial biopsy samples were collected from 22 YMA and 22 AMA women at day 5 of progesterone administration (P+5) undergoing endometrial receptivity testing using 57 common transcriptomic markers. Study samples (12 YMA and 12 AMA) underwent RNA sequencing and other samples (10 YMA and 10 AMA) were used for qPCR validation. FFPE tissue blocks were prepared for all study samples and hematoxylin/eosin-stained slides were evaluated by a clinical pathologist. Immunohistochemistry (IHC) analysis for p16^INK4a^, STC1, C2CD4B and LhS28 was performed in endometrial samples of both groups. Sequencing data were processed for quality control (QC), adaptor trimming and alignment to the human genome. Aligned reads were counted and analysed for differential expression (DE) between YMA and AMA groups. Computational deconvolution analysis was performed using single-cell RNA sequencing data from six endometrial cell populations. The figure is created with BioRender.com. **b**. The tissue computational deconvolution analysis. Fluidigm C1 single-cell gene expression dataset for six endometrial cell populations from Wang et al. (GSE111976) was used as a primary dataset for deconvolution analysis to estimate the proportions of six transcriptomically distinct cell types in endometrial tissue, and 10× dataset was used for validation. Briefly, read count matrices of distinct endometrial cell types were used to calculate the proportions of each cell type in the bulk endometrial samples using the dampened weighted least squares method (DWLS). DWLS is an estimation method for gene expression deconvolution, in which the cell-type composition of a bulk RNA-seq data set is computationally inferred. This method corrects common biases towards cell types that are characterized by highly expressed genes, to provide accurate detection across diverse cell types^50^.

**Fig.2.**
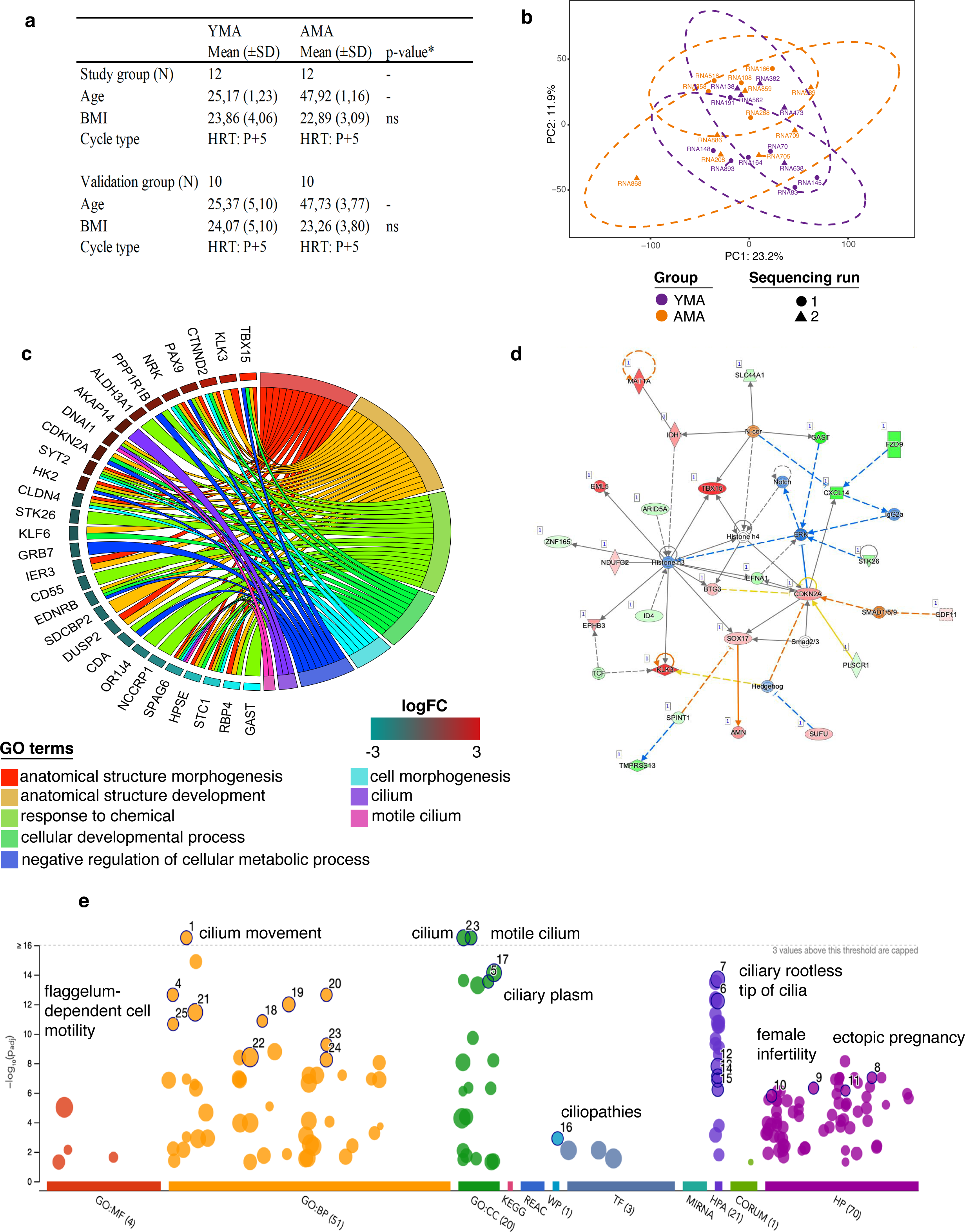
Genes dysregulated in AMA endometrium. **a.** Study participants. N – number of participants in each group, SD – standard deviation, HRT – hormonal replacement therapy, P+5 – day 5 of progesterone administration, ns – non-significant. *Unpaired Student t-test p-value. **b**. Principal component analysis of the study group samples based on RNA sequencing. **c**. Top DE genes and their biological functions. **d**. Molecular interaction network generated by Ingenuity Pathway Analysis (IPA). One of the top significant networks suggests the interaction between p16 (CDKN2A) senescence marker with Notch, ERK and Hedgehog signaling. **e**. Functional analysis of DE genes in AMA samples represents gene ontology (GO) terms significantly enriched in endometrium of AMA samples compared to YMA. BP – biological processes, CC – cellular component, HP – human phenotype, MF – molecular function, REAC – reactome, TF – transcription factor, HPA – human protein atlas, WP – WikiPathways.

After multiple testing corrections, 491 significantly differentially expressed genes (DEGs) (271 up-regulated, 220 down-regulated) were identified in the endometrium of the AMA versus YMA group (see Supplementary File 3). The expression rate changes were in range between -7-fold and 7-fold. The genes with the highest expression fold change were *GAST* (logFC = -2.82), *TBX15* (logFC = 2.81), *RBP4* (logFC = -2.62), *GP2* (logFC = 2.52), *ADGRF1* (logFC = -2.29), *STC1* (logFC = -2.26), *S100P* (logFC = -2.17), *HPSE* (logFC = -2.11), *DEPP1* (logFC = -2.08), *KLK3* (logFC = 2.04), *CXCL14* (logFC = -2.02) and *CTNND2* (logFC = 2.02). Most significant genes were *ZNF229* (p adj = 1.80×10^-8^), *C2CD4B* (p adj = 1.80×10^-8^), *TBX15* (p adj = 1.93×10^-7^), *SPAG6* (p adj = 3.56×10^-6^), *OR1J4* (p adj = 2.79×10^-6^), *TOR4A* (p adj = 2.79×10^-6^), *CLDN4* (p adj = 4.40×10^-6^), *GRB7* (p adj = 5.34×10^-6^), *C2CD4A* (p adj = 5.34×10^-6^), *RBP4* (p adj = 7.28×10^-6^), *PPP1R1B* (p adj = 1.28×10^-5^), and *RIMKLB* (p adj = 2.42×10^-5^) (Supplementary File 3).

Functional analysis of the genes differentially expressed in AMA endometrium showed that the most enriched gene ontology (GO) terms were general cellular and organismal developmental processes. These results are depicted in the Fig. 2c. Significantly enriched cellular components were cilium (GO:0005929; p adj = 0.013) and motile cilium (GO:0031514; p adj = 0.043). Additionally, the human phenotype ontology terms were abnormal respiratory motile cilium morphology (HP:0005938, p adj = 0.039) and abnormal respiratory epithelium morphology (HP:0012253, p adj = 0.039). All significant terms are presented in Supplementary File 4. IPA proposed five significant molecular networks. The most enriched networks were associated with cancer and general organismal functions, cellular (including immune cell) movement, endocrine system disorders (including metabolism disorders), cell morphology, and development (see Supplementary Table 2 and Supplementary Fig. 1-5). The most enriched upstream regulator was TP63 (p = 3.4×10^-6^, see Supplementary Table 2), a transcription factor that regulates epithelial cell fate through interactions with Sonic Hedgehog (Shh), Wnt, and Notch pathways^13^. The molecular interactions network with the highest score was associated with cellular movement, development, and cancer. DEGs are directly or indirectly associated with the cell cycle regulator molecule cyclin-dependent kinase 2A (CDKN2A), better known as p16^INK4a/ARF^ locus, which encodes a cellular senescence marker and presented interactions with Shh and Notch pathways (Fig. 2d, Supplementary File 4). Functional annotation with g:Profiler confirmed that most significantly enriched biological processes were associated with cilial movement and most enriched cellular components were cilium and motile cilium (Fig. 2e, Supplementary File 4).

When examining DEGs for their putative role in endometrial functioning, it was found that many upregulated genes were associated with decidualization (*ALDH3A1, KCNE1*), endometrial receptivity (*GALNT12, TMED6, CLDN4, CD55*), cell cycle control (*CDKN2A*), and insulin signaling (*TMED6*), while downregulated genes included sugar metabolism and inflammation (*C2CD4A, C2CD4B, NFKB*), cellular movement and invasion (*SPAG6, HPSE, GRB7*), and ovarian hormone signaling (*STC1, GRB7, ALDH3A1*). When compared to previously published genes associated with endometrial receptivity^8–11^, 72 genes out of 491 genes DE in AMA samples were associated with endometrial receptivity in natural cycles, and 18 genes were associated with hormonal replacement therapy (see Supplementary File 6).

### The expression of stanniocalcin-1, p16^INK4a^ and C2CD4B in the endometrium of AMA group

To localize the products of DE genes in AMA endometrium, immunohistochemical analysis was performed on decidualization-associated protein STC1, cellular aging marker p16^INK4a^ and receptivity marker C2CD4B (also known as NLF2) in 4 YMA and 4 AMA samples. The analysis of protein staining intensity showed that p16^INK4a^ staining intensity is significantly higher in the luminal and glandular epithelial cells of AMA samples (p<0.05, Wilcoxon rank-sum test, two-sided) (Fig. 3a,b,j). Immunohistochemical analysis showed downregulation of STC1 protein in all three cellular compartments, however it was not significant between two groups (Fig. 3d,e,j). C2CD4B was upregulated in luminal epithelial and stromal cells of YMA samples, potentially implying compromised receptivity with age (Fig. 3g,h,j). All three analyzed protein markers showed a higher degree of variation in the AMA group than in the YMA group, indicating a possible age-related endometrial tissue heterogeneity. Real-time quantitative PCR experiments using validation group samples confirmed the expression changes of several DE genes in AMA group *ALDH3A1, EML5, SPAG6, STC1, PPP1R1B* and *TBX15* in AMA group (Fig. 3d). Significant changes (p-value < 0.05) are indicated by an asterisk.

**Fig.3.**
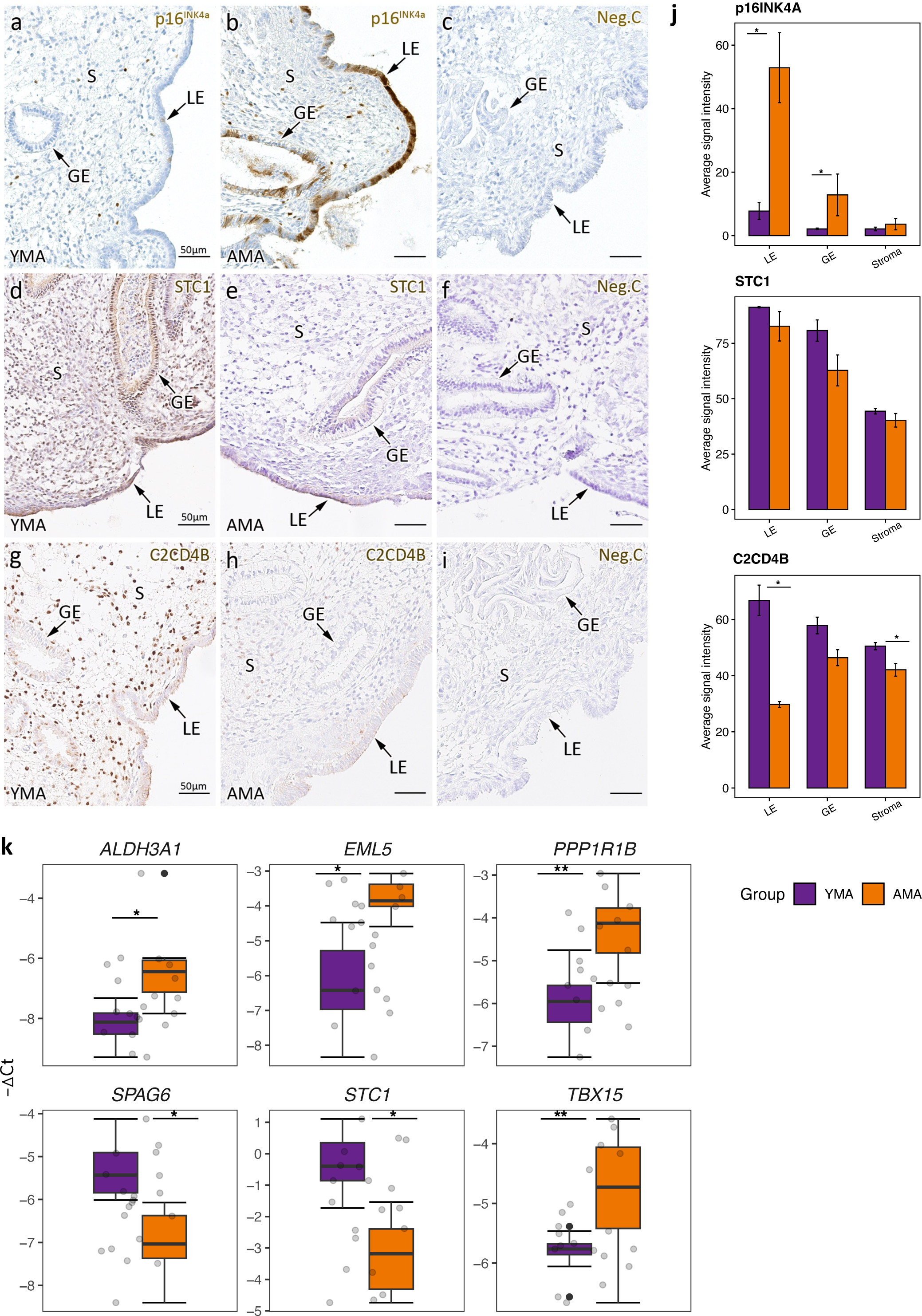
Immunohistochemistry analysis of proteins of selected DE genes and validation of RNA-seq results. **a-b**. The staining of p16^INK4a^ senescence protein in YMA and AMA samples, respectively. **d-e**. The staining of decidualization-associated protein stanniocalcin-1 in YMA and AMA samples, respectively. **g-h**. The staining of receptivity and reproductive outcome-associated protein C2CD4B (also known as NLF2) in YMA and AMA samples, respectively. **c, f, i.** negative controls for each experiment. LE – luminal epithelium, GE – glandular epithelium, S – stroma. **j.** p16^INK4a^, STC1 and C2CD4B protein expression in endometrial cellular compartments, identified by IHC from YMA patients (n = 4) and AMA patients (n = 4). \**P<0.05*; Wilcoxon rank-sum test. Bars represent means with SEM. **k**. qPCR validation of DE genes in AMA group identified by RNA-seq. \**P<0.05*; Wilcoxon rank-sum test.

### Proportions of endometrial cell populations in YMA and AMA groups

Histological evaluation of study group samples according to Noyes criteria confirmed that all endometrial samples were in the secretory phase and complied with their endometrial receptivity status (Pearson positive correlation p=0.001).

Deconvolution analysis using the C1 single-cell dataset of six endometrial cell types from GSE111976 showed a significantly higher proportion of ciliated epithelial cells in AMA samples compared to YMA samples (proportion, 0.01 ± 0.005 versus 0.004 ± 0.002; p<0.05, Wilcoxon rank-sum test, two-sided; Fig. 4a,b). The same analysis was performed with the 10× single-cell dataset of eight endometrial cell types as a reference. It also revealed that ciliated epithelial cells are more abundant in AMA samples as compared to YMA counterparts (proportion, 0.006 ± 0.009 versus 0.0009 ± 0.002; Fig. 4c,d), although the difference was not statistically significant. This could be due to the different sampling or library preparation techniques between two single-cell references. When the samples were ordered by age, it became clear that the AMA group samples had a higher variation in the proportions of ciliated cells compared to the YMA group (Fig. 4e).

**Fig.4.**
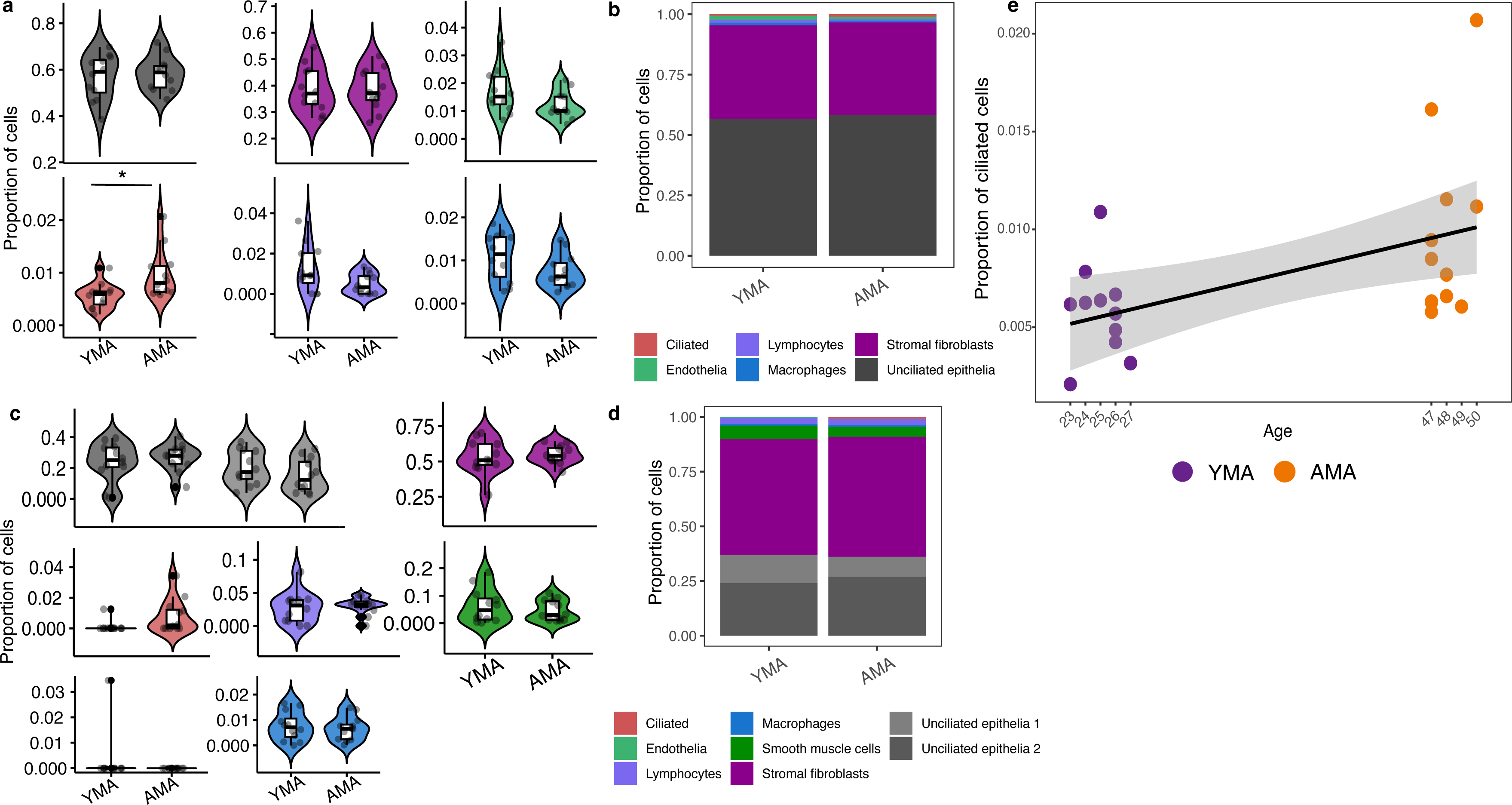
Cell type proportions per age group determined by deconvolution analysis in 24 samples. All cell types have a color associated as shown in the legend. **a.** Violin plots show the changes in cellular proportion between young and advanced maternal age groups for six cell types based on the C1 single-cell RNA-sequencing dataset. The difference in ciliated cells between young and advanced maternal age groups is statistically significant, indicated by * (two-sided Wilcoxon test with Bonferroni correction, p-value < 0.05). The boxplots show the interquartile range (box limits) and median (center black line) of cellular proportion levels. **b**. Stacked bar plots depict percentages of six cell types in young and advanced maternal age groups based on the C1 single-cell RNA-sequencing dataset. **c.** Violin plots show the changes in cellular proportion between young and advanced maternal age groups for eight cell types based on the 10× single-cell RNA-sequencing dataset. The boxplots show the interquartile range (box limits) and median (center black line) of cellular proportion levels. **d.** Stacked bar plots show percentages of eight cell types in young and advanced maternal age groups based on the 10× single-cell RNA-sequencing dataset. **e**. Proportions of endometrial multiciliated epithelial cells aligned according to woman’s age.

### The regulation of AMA genes across menstrual cycle

To test whether the DE genes in AMA may have an impact on the window of implantation (WOI), we analyzed the expression of AMA genes across the menstrual cycle in six endometrial cell types. *In silico* analysis using transcriptomic data from GSE111976 dataset was performed. As a result, it appeared that *C2CD4B, LYPD3, MFSD4A, RBP4, CXCL14, GAST, HPSE* and *TMEM92* were up, and *SPDEF* was down-regulated in endometrial secretory epithelial cells during mid-secretory phase (Supplementary File 5). Single-cell data also showed that *AKAP14, c11orf97, CAPSL, DNAI1, SPAG6* and *WDR49* were highly expressed in ciliated endometrial cells across the menstrual cycle, compared to other cell types. However, their expression in the whole tissue is modest due to its small number of ciliated cells (Supplementary File 3).

### Immunohistochemical analysis of multiciliated cells in AMA endometrium

An immunohistochemical analysis of cilia basal bodies (BB) using anti-LhS28 antibody was conducted to determine if AMA samples had more ciliated cells than YMA samples. The stained BBs of multiciliated epithelial cells were observed as halfmoon-shaped organelles near the surface of epithelial cells, with motile cilia often visible on top (Fig. 5a-c). The number of positively stained BBs was counted per area of luminal epithelium (LE) in both YMA and AMA groups during image analysis (Fig. 5e). The average BB count per LE area was higher in the AMA group compared to the YMA group (3.6*10^-3^ (SD ±0.8*10^-3^) vs 2.0*10^-3^ (SD ±0.9*10^-3^), respectively; Wilcoxon unpaired sample p-value 0.028), which supported the results of deconvolution. The AMA DE genes that regulate cilial development and function are shown in Fig. 5f.

**Fig.5.**
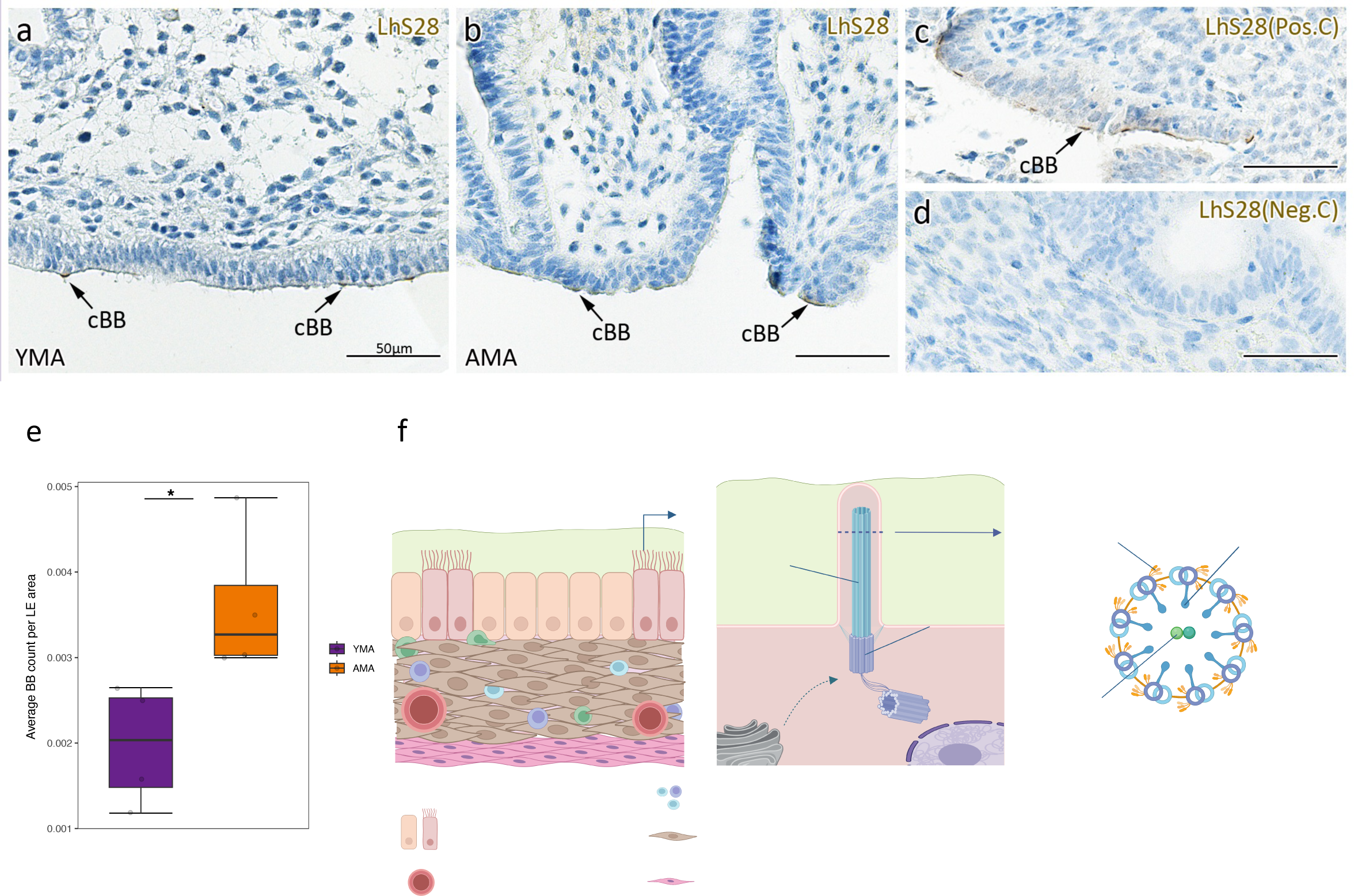
The analysis of multiciliated cells in YMA and AMA endometrium. **a-d**. Basal body staining using LhS28 antibody in endometrial tissue: YMA, AMA, day 16 of natural cycle (positive control) and negative control, respectively. **e.** Relative basal body (BB) count per luminal epithelium (LE) area calculated based on LhS28 staining in human endometrium. *significant, independent two-sample Wilcoxon test p<0.05. **f**. Structure of endometrial multiciliated cells and functions of motile cilia affected by AMA. The gene names represent the genes DE in AMA endometrium and associated with motile cilia, based on previously published evidence. The figure is created with BioRender.com

## Discussion

This retrospective study compared the transcriptome profiles of endometrial biopsy samples during WOI from young and very advanced reproductive age groups. Compared to other endometrial aging studies, we analysed participants that underwent similar endometrial preparation protocols before IVF that minimizes physiological bias. Our study provides considerable evidence that a woman’s age affects the development of endometrial tissue and its function. For example, we discovered that *TBX15* was the most highly upregulated gene in the advanced maternal age group, while *RBP4* was the most downregulated. *TBX15* is a gene expressed by epithelial cancer cells and associated with epithelial to mesenchymal transition (EMT)^12^. The upregulation of *TBX15* reduces apoptosis through NF-κB^13^. EMT is normally induced by embryonic signals and helps endometrial luminal cells to acquire the characteristics of motile epithelial cells, accommodating incoming embryo during implantation^14^. Lower levels of *RBP4* were shown to reduce the invasion ability of trophoblast cells *in vitro* and lower serum levels of *RBP4* have been associated with higher risk of pre-eclampsia^15^. This paracrine molecule is secreted into the endometrium to inactivate the excess oestradiol activity during decidualization^16^. Also, *STC1*, a well-studied progesterone-responsive gene that defends cells from oxidative stress, is upregulated in receptive endometrium and promotes decidualization of stromal cells. Its dysregulation in eutopic endometrium suggests involvement in the pathogenesis of decidualization events^17, 18^.

In addition, both *C2CD4A* and *C2CD4B* were downregulated in the AMA group. Their gene products are involved in the regulation of vascular permeability during acute inflammatory response. *C2CD4B* takes part in the signalling pathway causing changes in cell architecture and adhesion in endothelial cell inflammation^19^. It is also strongly expressed in the mid-secretory endometrium, emphasizing its role in endometrium remodeling during the WOI, while its downregulation is associated with recurrent implantation failure^20, 21^. Moreover, *C2CD4A/C2CD4B* locus regulates the susceptibility to Type 2 diabetes, a disease characterized by complex metabolic and inflammatory processes, strongly associated with age^19^. The downregulation of *C2CD4B* and *STC1* in AMA samples suggests a direct negative effect of maternal age on endometrial receptivity and implantation success.

The pathways most affected by age were cellular remodeling, cilial movement and cell migration, and immune response. Interestingly, some of the identified DE genes (e.g., *CDKN2A, ALDH3A1, ACE, EDNRB*) have been previously linked to aging. Isoforms of *CDKN2A* encode cyclin-dependent kinases p16^INK4a^ and p14^ARF^ that participate in the estrogen-responsive cell cycle regulation by hindering G_1_ phase progression and promoting apoptosis. P16^INK4a^, also known as p16, common aging biomarker^22^, is expressed in both epithelial and stromal endometrial cells. Immunohistochemical analysis showed a significant difference in p16 staining between AMA and YMA luminal epithelia with higher expression in AMA group. Although it was suggested that a higher amount of p16-positive epithelial cells was required for a supportive environment during the window of implantation, it was also previously shown that p16-staining in endometrial epithelial cells correlated positively with women’s age^22, 23^. The involvement of p16 in the cellular aging mechanism is associated with its role in cell cycle progression. The transition from G1 to S phase requires the activation of the cyclin-dependent kinases CDK4 and CDK6, which inactivate retinoblastoma (Rb) tumor suppressor through phosphorylation^24^. Rb phosphorylation promotes the expression of genes necessary for S-phase entry. p16^INK4a^ is a cell cycle regulator that inhibits the activation of CDK4/6, resulting in cell cycle arrest at the G1 phase. Senescent cells cease proliferation and secrete factors that initiate the senescence-associated secretory phenotype (SASP), which includes events that recruit immune cells and clear senescent cells for tissue regeneration^25, 26^. However, if the elimination of senescent cells does not occur, senescent cells accumulate and lead to cancer or tissue aging^27, 28^. SASP also affects epithelial barrier integrity through disorganized tight junction proteins that make tissues more susceptible to pathogens. Our molecular network analysis suggested the interaction of p16^INK4a^ (*CDKN2A*) with Notch transmembrane receptor signaling, which also regulates cell cycle progression (Fig. 2d). Activated Notch receptors lead to dephosphorylation of Rb, which produces flat and enlarged cell morphology, and senescence-associated beta-galactosidase activity^29^. In fact, it was proposed that activated Notch induces the apoptosis or cell fate change in multiciliated cells in vertebrate models^30^, and can act both as oncogene and tumor suppressor, but its action in endometrial multiciliated cells is largely understudied. It was shown that senescence-messaging secretome can trigger overexpression of p16^INK4a^ in endometrial stem cells by activating the paracrine stress-induced mechanism of premature senescence^31^.

Our results suggest the involvement of p16-associated cellular senescence and the suppression of metabolic and inflammatory processes which are essential for preparing the endometrium for embryo transfer. Although it is unclear how p16 accumulation affects human endometrial epithelium, it has been shown that higher amounts of p16 mRNA correlate with greater primary cilia volume in the stem cells of cervical epithelium in mice^32^. In humans, p16 overexpression is associated with endometrial tubal metaplasia, characterized by a higher amount of ciliated cells than endometrium, and is considered a specific marker for dysplastic and neoplastic epithelial cells^33, 34^. On the contrary, higher amounts of p16-stained cells in both glandular and luminal compartments were associated with higher live birth rates and lower miscarriage rates in women undergoing IVF^23^, although they did not discriminate for ciliated cells. Our experiments also do not provide evidence that p16-associated senescence affects cilial development. Instead, it is more likely that changes in the development of ciliated epithelial cells can affect stem cell recruitment and impact the overall epithelial regeneration process.

Functional analysis of DE genes revealed that the pathways associated with motile cilia formation and ciliopathies were highly significant in AMA endometrium (Fig.2e). In total, 71 out of 491 DE genes appeared to be involved in cilial structures and function (see Supplementary File 3), with almost all of them upregulated in AMA samples. Interestingly, the deconvolution analysis also showed a significantly higher proportion of ciliated epithelial cells in the AMA group (Fig. 4a-e). Cilia are microtubule-based cellular structures predominantly used for sensing, with the primary cilium having 9+0 axonemal structure and the motile cilium having an additional central pair of microtubules (9+2 axonemal structure) and accessory structures such as dynein arms that allow for movement. In addition to sensory functions motile cilia are necessary for liquid moving along the epithelial surface. In female reproductive tract, motile cilia help move mucus and support an embryo. Ciliogenesis is driven, although not exclusively, by estrogen and through inhibition of Notch signaling^35, 36^. Multiciliated epithelial cells are unable to divide due to the use of centrioles in cilia formation^30^. Rising estrogen levels during the menstrual cycle promote ciliated cell proliferation and differentiation from epithelial progenitor cells, and they reach their peak by day 15-16 of the menstrual cycle. At that point, progesterone produced by the corpus luteum opposes estrogen action and reduces the amount of multiciliated cells significantly by the time implantation should take place^37, 38^. This cyclic event highlights the importance of multiciliated cells in regulation of endometrial receptivity, including both their sufficient proliferation and programmed decrease. Our results showed the dysregulation of Notch-associated genes in the AMA group (e.g. *DNAI1, DNAH3, DNAH6, DNA9, DNAH11, DNAH12, GNMC*) and the possible opposing effect of CDKN2A on Notch signaling through inhibition of ERK (Fig. 2d). Together with the downregulation of progesterone regulated genes *STC1, CLDN4, ARID5B, VEGFA* and *C2CD4B* genes, higher proportion of multiciliated cells may also be the signs of suboptimal progesterone action in AMA endometrium. Normally, progesterone levels gradually decrease over years before the last flow and this may impact the activity of progesterone-dependent signaling^39^. Although synthetic progesterone is administered to activate progesterone receptors in the endometrium during HRT, transcriptomic studies showed the differences between hormonally induced and natural human mid-secretory endometrium^11^. It is unclear whether aging may affect the way progesterone acts in human endometrium, but there are animal studies that demonstrated the possible effect of aging on progesterone sensitivity^40^.

Previous studies have shown that multiciliated cells are a distinct cell subtype within endometrial epithelium and have a specific transcriptomic profile^41, 42^. Our results suggest that the expression of many genes involved in motile cilia development (Supplementary File 2) were altered in AMA, such as motile cilia development genes *ARMC6, FAM183A, VWA3A*, dynein axonemal chain genes *DNAI1, DNAH3, DNAH6, DNAH9, DNAH11, DNAH12*^42^, radial spoke head component proteins genes *RSPH1* and *RSPH4A*, outer dynein docking complex subunit 4 gene *TTC25*, and a number of cilia-associated genes, such as *PIFO, CFAP43, CFAP45, CFAP52, CFAP57, CFAP58, CFAP61, CFAP65, CFAP73, CFAP77, CFAP100, CFAP157, CFAP161, c1orf189, CATIP* and *SNTN*, present in both primary and motile cilia^43^. The cilia-associated DE genes were present in all structural parts of cilia and were responsible for both cilial development and function (Fig. 5f, Supplementary File 3). By the way, all cilia-associated genes, except for *SPAG6*, were upregulated in the whole-tissue samples of AMA group. This raises the question of whether these changes are due solely to the increased proportion of ciliated cells in the AMA samples or if multiciliated cells also exhibit unique transcriptomic changes in AMA samples. *SPAG6* is an estrogen-induced gene that promotes cellular migration and is upregulated in progressive reproductive cancers^35, 44^. On the contrary, a strong reduction in *SPAG6* mRNA is a sign of EMT in certain cancer types. This together with the highly significant upregulation of *TBX15* suggest the signs of altered EMT in AMA samples. Previously, age was reported to be negatively correlated with ciliary beat frequency, and associated with cilial structure abnormalities^45^. This supports the hypothesis that not only the number of ciliated cells is affected by age, but the development of cilia itself.

Our findings agree with the previous research conducted by the Diaz-Gimeno group, in which they used *in silico* analysis to identify DE genes related to ciliogenesis^7^. However, only few genes (such as *DNAI1*, DNAH9 and *CCDC39*) overlap between our study and theirs, which may be attributed to the differences in analysis methods and age thresholds. In contrast to their study, which used Affymetrix array transcripts, our study utilized RNA-seq analysis with a different annotation. Our results indicate that genes associated with motile cilia were not very highly expressed in the whole tissue sample (baseMean range between 23-533, see Supplementary File 3), implying that many molecules in the same molecular cascade could have been missed during whole tissue analysis. Nevertheless, we found that age significantly affects the molecular profile of the endometrium, resulting in notable changes in epithelial cell regeneration. It is uncertain whether these changes directly impact endometrial receptivity, but our findings provide compelling evidence that age affects many processes associated with endometrial development.

Along with cilia-associated genes and general cell cycle-regulating genes, we identified a set of DEGs previously described in association with endometrial receptivity. Among those are several common endometrial receptivity-associated markers, such as *CXCL14, FOSL2, S100P, CLDN4, ID4, SLC15A1, SLC1A1, ARID5B, MRPS2, LAMB3, IER3, EFNA1, EDNRB, DDX52, HPSE, BCL6, GAST, CDA* and *HABP2*^46–48^. Significantly downregulated *C2CD4B* was also reported as an endometrial receptivity gene. It is up-regulated during the mid-secretory phase in human endometrium and is shown to be lower in HRT patients compared to natural cycles^49^. One of the most strongly expressed genes *IDH1* (baseMean 21 840.94) encoding isocitrate dehydrogenase 1, highly expressed in all types of epithelial cells, was 2-fold up-regulated in AMA samples (p adj = 6.15*10-3) (Supplementary File 3) is also a hub gene in RIF^50^. Also, *SPDEF* encoding SAM pointed domain containing ETS transcription factor, is one of the genes upregulated and *HABP2*, hyaluronan binding protein 2, is downregulated in patients with recurrent implantation failure^11^. *SPDEF* is a vascular development protein essential in biochemical pregnancy establishment^51^. Processes of vascularization may be disrupted in AMA endometrium, as genes such as *S100P, GAST, ID4*, and *EFNA1* which need to be upregulated for normal vascular development, are downregulated in older women^51^. The disturbance of estrogen signaling by AMA is supported by the changes in direct estrogen-responsive genes, such as *GRB7, CD55*, and *CLDN4*. *GRB7* regulates integrin signaling pathway and cell migration by interacting with focal adhesion kinases (FAK) and phosphatidylinositol 3 kinase (PI3K)^52^. Integrin signaling and FAK are also inhibited by the upregulation of previously mentioned *SPDEF*, as suggested by IPA molecular network 2 (see Supplementary File 4). Heparanase (HPSE) was proposed as a biochemical pregnancy biomarker and its upregulation in the extracellular matrix is essential to improve embryo-endometrial adhesion. During healthy pregnancy, it also supports angiogenesis and placentation^53–55^. Lower heparanase mRNA, as observed in AMA samples, is associated with the risk of consecutive miscarriages^53^. The dysregulation of these genes suggests that endometrial aging affects tissue vascularisation, cellular migration, and adhesion, involved in the establishment of endometrial receptivity.

To sum up, our study provides considerable evidence that a woman’s age affects the development of endometrial tissue and its function. The current study is the first to show that older women have a significantly higher proportion of ciliated epithelial cells in their endometrium. However, it is not clear whether this shift is caused by a larger production of ciliated cells or their insufficient decrease under the influence of luteal phase hormones. Excess of estrogen and lack of progesterone activity may both produce a larger amount of multiciliated cells. Gene and protein expression experiments support the idea of a woman’s age effect on the way epithelial cells respond to progesterone. The development of epithelial multiciliated cells is strongly associated with cell cycle and can be implicated by cellular senescence. Here we speculate that the accumulation of senescent epithelial cells may also lead to the shift in the proportions of epithelial cell subpopulations and point to mild tubal metaplasia in advanced reproductive age women. It is not quite clear whether this shift is enough to cause implantation failure or if it adds to the other factors compromising endometrial receptivity, such as mild inflammation and altered sugar-lipid metabolism. Also, to understand whether the differences in gene expression of whole tissue samples uprise from the changes in the cell proportions or if there are significant differences in the development and metabolism of specific cell types that occur with age, a prospective study utilizing cell type-specific gene expression of different cell types from AMA endometrium should be performed. Given the diversity observed in endometrial samples from women over 45, it is evident that aging does not occur at a specific time point and is peculiar for every individual. Therefore, a more accurate molecular definition of endometrial aging is required to address this issue.

Also, our study is not without limitations. Although our data show that AMA women have a higher proportion of ciliated epithelial cells in the endometrium during the WOI than younger women, this is a retrospective study, and our clinical data is limited. We do not have the blood hormone levels measured for the participants, as the patients have undergone HRT, the hormones are nor measured during routine endometrial receptivity screening. The endometrial pathology data may also be not fully inclusive. To overcome this, a prospective clinical study involving thorough participant selection plan and actual counting of multiciliated epithelial cells in association with the pregnancy outcome should be conducted to test whether the abundance of multiciliated cells could serve as a robust diagnostic marker for infertility management in women of advanced maternal age.

## Materials and Methods

### Patients and samples

Anonymized endometrial biopsies were obtained from 44 women with an assessed endometrial receptivity status based on 57 common endometrial receptivity markers (as described in Meltsov et al^56^) in hormonally induced cycles and all samples had the estimated receptivity score between 70 and 110, identified as ‘receptive’. The study group consisted of 12 women aged 20-27 (young maternal age group, YMA) and 12 women aged 47-50 (advanced maternal age group, AMA), and the validation group consisted of 10 YMA and 10 AMA samples. All women were infertility patients planning frozen embryo transfer in HRT cycle. The only inclusion criterion was the absence of reported uterine pathologies. According to available clinical data, the progesterone was administered orally, vaginally, or by injection. The first day of progesterone administration was considered as day zero (P+0). Endometrial receptivity screening test result was used as a reference to evaluate the progesterone efficiency in endometrial tissue. An endometrial biopsy was taken on day 5 of progesterone administration (P+5). All endometrial tissue biopsies were collected using a Pipelle catheter (Laboratoire CCD, France) and immediately placed into RNAlater (Ambion, USA). The overview of the study design is presented in Fig.1.

### RNA sequencing of endometrial samples

#### Library preparation and sequencing

Total RNA was extracted from all samples using miRNeasy kit (Qiagen, cat.no. 217084), followed by DNase treatment (DNase Free, Life Technologies, cat.no. EN0521), according to the manufacturer’s protocol. RNA integrity was assessed with Qubit RNA IQ Assay kit (Thermo Fisher, cat.no. Q33221). cDNA libraries were randomly assigned into two sequencing batches and prepared using the TruSeq Stranded mRNA Sample Preparation Guide (Illumina, cat.no. 20020595). The libraries were then pooled and single-read 75bp sequencing was performed on NextSeq500 (Illumina).

#### Read mapping and quality control

Reads were mapped to the *Homo sapiens* reference genome (UCSC release GRCh37/hg19), using *STAR* (v.2.7.9) in “Basic” two-pass mode and in “GeneCounts” quantification mode. FastQC (v.0.11.9) and samtools (v.1.14) were used to extract quality control metrics. All the samples had more than 20M reads aligned to the genome with the alignment percentage of > 85%. The quality control metrics and the alignment statistics were summarized in the HTML report using MultiQC (v.1.12) (see Supplementary File 1).

The mapped reads were estimated at gene level using the Ensembl database for annotation. The raw expression count matrix was produced using *FeatureCounts* (v.2.0.1). Batch effect of the sequencing run was corrected for using the ‘Combat-Seq2’ method from the *sva* R package (v.3.42.0) as it uses a negative binomial regression model, suitable for modeling the characteristics of bulk RNA-seq count data. Subsequently, genes in which the number of mapped reads was <5 in more than 75% of samples were excluded from the further analysis. General sequencing results statistics and sample clustering is presented in Supplementary File 1.

#### Differential expressed genes detection

To identify the differentially expressed genes (DEGs), the matrix of batch effect-adjusted counts was supplied to *DESeq2* (v.1.34.0) using a filter criterion of false discovery rate (FDR) < 0.05. The receptivity score was used as a covariate in the design matrix. Since the expected variance of RNA-seq counts increases with the mean, the ‘variance stabilizing transformation (vst)’ function from *DESeq2* was used to generate a matrix of transformed counts for sample visualization. To check endometrial gene expression in relation with age, the top 20 most significant genes were sorted by expression log fold-change (logFC) and clustered hierarchically using the vst-modified counts. This was plotted in heatmaps with accompanying dendrograms using the *pheatmap* package in R (v.1.0.12). The heatmap and the dendrogram are presented in Supplementary File 1.

#### Functional analysis of DE genes

Enrichment analyses for Gene Ontology (GO) terms and biological pathways were done using g:Profiler (v.0.2.1, updated June 14,2022)^8^ and Ingenuity Pathway Analysis (IPA) (Qiagen) tool using all significant differentially expressed genes (DEGs) identified between AMA and YMA groups. For graphical representation of functional analysis data GO chord plot was generated using the *GOplot* (v1.0.2) package in R. Networks of significant DEGs were then algorithmically generated based on their connectivity in IPA. The DE genes were then compared with a set of endometrial receptivity genes regulated during WOI of healthy women, reported in publications analyzing RNA-seq of human endometrial mid-secretory samples^8–10^ and women undergoing HRT^11^. UMAP and PCA plots were generated using *embed* (v1.1.0) and *recipes* (v1.0.4) packages, Venn diagrams with *ggvenn* package (v0.1.10) and ggplot for boxplots (v3.4.0).

#### Power calculation

Power analysis for RNA-seq experiments is essential for determining the appropriate sample size required to achieve the desired level of statistical power. This ensures that the experiment is adequately designed to detect differential expression (DE) between two groups while minimizing the risk of false positive and negative results. Performing power analysis for RNA-seq experiments is challenging due to the complexities associated with the nature of the data and the methods used for analysis. Thus, analytical solutions for power calculations in RNA-seq experiments are not feasible. Instead, the PROPER (PROspective Power Evaluation for RNAseq) R package^57^ provided a simulation-based method to compute power-sample size relationships. With this method it was computed that given the sample size 12, significance level (p adj) 0.05 and the effect size (logFC) 0.3, the power estimate was 85%, whereas for the genes with logFC at least 1, the power was 99%.

### Quantitative real-time PCR (qRT-PCR) validation of sequencing results

Total RNA was extracted from the endometrial samples of the validation group (YMA n = 10, AMA n = 10) and DNase-treated as described above. cDNA was synthesized using Maxima First Strand kit (Thermo Fisher, USA) and qRT-PCR reactions were performed using HOT FIREPol EvaGreen qPCR Mix (Solis BioDyne, Estonia) and the standard protocol as previously described in Suhorutshenko et al.^8^. Oligonucleotide primers for *ALDH3A1, SPAG6, STC1, PPP1R1B, EML5* and *C2CD4B* are presented in the Supplementary File 2. All reactions were performed in triplicate. Expression values were normalized to the endogenous control genes *TBP* and *GAPDH*. Data were analyzed using 7500 Software v2.0.5 (Applied Biosystems). Differences in gene expression levels between AMA and YMA samples were estimated using comparative Ct (2^-ΔΔCt^) method^58^. Unpaired two-sample Wilcoxon test was applied to estimate significance level in R.

### Histological analysis

For histological analysis of study group samples, 24 microscope slides with tissue sections (4 μm, 3 sections per slide) were prepared at the Tartu University Hospital Pathology Department (Tartu, Estonia) from formalin-fixed paraffin-embedded (FFPE) endometrial tissue sections using standard hematoxylin and eosin staining protocol^59^. The slides were scanned at East Tallinn Central Hospital, using a 3DHistech Pannoramic Flash III 250 scanner (3DHistech, Budapest, Hungary) at a 20× magnification. Evaluation of cyclic endometrial changes was performed according to Noyes criteria^60^ by a clinical pathologist (A.M.).

### Immunohistochemistry

To evaluate the expression of cell cycle progression and cellular senescence protein p16^INK4a^, decidualization-associated protein STC1 and glucose metabolism-associated protein C2CD4B, the immunohistochemistry (IHC) analysis was performed. For IHC, four samples from both YMA and AMA groups were selected at random, with a key consideration being that the average receptivity score should not differ significantly between the groups. FFPE tissue sections on slides were processed according to standard protocol of Master Polymer Plus Detection System IHC Kit (MAD-000237QK-10, Master Diagnostica, Spain) was used. Incubation with primary antibody was carried out overnight in a humidity chamber at 4L. Further steps were carried out as per manufacturer’s protocol. Chromogenic reaction was developed for 30 seconds and stopped thereafter. Cell nuclei were counterstained with Mayer’s haematoxylin solution for 15 seconds, then the slides were washed for 10 minutes with tap water. Slides were dehydrated through graded ethanol and xylene solutions and mounted with Leica CV mount (Leica Biosystems, US).

Following primary antibodies were used: rabbit polyclonal IgG against human STC-1 (Atlas Antibodies, Stockholm, Sweden) dilution was 1:200 (in 1% BSA/PBS), anti-p16^INK4a^, mouse monoclonal IgG against human CDKN2A was ready-to-use antibody solution (Master Diagnostica, Spain), and anti-C2CD4B (Sigma-Aldrich, Saint Louis, USA). For ciliated cells visualization, IHC staining of cilia basal bodies was performed using anti-LhS28 antibody (Santa Cruz Biotechnology, Germany) with dilution 1:200. As a positive control for LhS28 staining, additional endometrial biopsy taken on day 15 of natural menstrual cycle from fertile young maternal age woman was used. All antibodies used in this study are listed in Supplementary File 2.

### Image and data analysis

Slides were scanned using Leica SCN 400 Slide Scanner (Leica Biosystems, US) with a maximum of 20× magnification objective. Semi-quantitative analysis of three different areas of scanned sections was conducted with ImageJ package Fiji (v1.52e)^61^. Briefly, the intensity of DAB signal was measured separately for stromal, glandular epithelium and surface epithelium components of the endometrium. The relative DAB intensity was calculated using the formula: f = 255 - i, where “f” is relative DAB intensity and “i” is mean DAB intensity obtained from the software ranging from 0 (zero – deep brown, highest expression), to 255 (total white)^62^. The Wilcoxon Mann-Whitney test was performed to determine statistical significance. GraphPad Prism 6 (San Diego, CA, USA) and ImageJ software (Fiji package) were used for data quantification and analysis. Basal body counting was performed blinded by two independent specialists using anti-LhS28-stained endometrial tissue slides with 40× magnification and the average amounts of ciliated cells exhibiting basal bodies between two measurements were estimated. Unpaired sample Student’s t-test was applied to test statistical significance between the two groups.

### Deconvolution

Two single cell transcriptome datasets of endometrial biopsies – C1 dataset (library preparation method: Fluidigm; n cells = 2148) and 10× dataset (library preparation method: 10× Chromium Next GEM; n cells = 71032) – and their respective cell type labels were obtained from NCBI’s Gene Expression Omnibus (accession code GSE111976)^41^. The dataset was chosen based on the next criteria: completeness, annotation, sample size and availability of supportive information. This dataset is currently the most comprehensive and well-annotated single-cell dataset for the endometrial cell types. For the menstrual cycle phase concordance with the bulk RNA-seq samples, single-cell datasets were subset to only include the samples corresponding to the mid-secretory phase. It resulted in 546-cell and 46878-cell count matrices for the C1 and 10× datasets, respectively. These matrices of distinct endometrial cell types were used to calculate the proportions of each cell type in the bulk endometrial samples using the dampened weighted least squares method from *DWLS* R package (v.0.1.0). Dampened weighted least squares method was used as it shows high rare cell-type detection accuracy and best performance among the deconvolution methods that use single-cell RNA-seq data as input^63^. To compute the significance of differences in the proportions of each endometrial cell type in young maternal age versus advanced maternal age (YMA and AMA) samples, a non-parametric Mann-Whitney *U* test (two sides) was used, and Bonferroni correction was applied. To visualize whether the proportion of multiciliated epithelial cells correlates with a woman’s age, the samples were ordered according to age. Violin, bar, and scatter plots were produced using the R package ggplot2 (v.3.4.0).

### Expression of the bulk DEGs on the single-cell data

The expression of 57 most significantly expressed protein-coding DEGs from the bulk data was visualized on the single-cell C1 dataset, obtained from NCBI’s Gene Expression Omnibus (accession code GSE111976), which covers the entire menstrual cycle. The raw single-cell counts were normalized using the shifted_log_transform() function from transformGamPoi R package (v.1.0.0).

## Code availability

All code used for the analysis is available at https://github.com/darinaobukhova/endo_ageing

## Ethics statement

We confirm that our research complies with all relevant ethical regulations. The research was approved by the Ethics Committee of the University of Tartu (Ethics Approval No. 340-12).

## Data Availability

All data produced in the study are available upon reasonable request to the authors

## Acknowledgements

This research was funded by the Estonian Research Council (grant PRG1076), Horizon 2020 innovation grant (ERIN, grant no. EU952516), Enterprise Estonia (grant no EU48695), MSCA-RISE-2020 project TRENDO (grant no 101008193) and EU 874867 project HUTER. Authors declare no competing interests.

## Supplementary Material

**Supplementary File 1**. Extended results of RNA-seq analysis. RNA-seq statistics, a dendrogram of the samples, and a heatmap of AMA genes.

**Supplementary File 2**. Primers and antibodies used for the expression analysis of AMA genes.

**Supplementary File 3**. Genes differentially expressed in AMA samples, including the cilia-associated genes.

**Supplementary File 4.** Functional analysis of DE genes in AMA. IPA and g:Profiler results.

**Supplementary File 5.** The expression of selected WOI-associated AMA genes in different cell types across the menstrual cycle. Dots represent samples from the single-cell sequencing dataset GSE111976 used as a reference in the study, colours represent the subsequent cell types. **Page 1**. Gene differentially expressed in different endometrial cell types across the menstrual cycle. **Page 2**. Genes with a significant change in their expression rate during WOI, corresponding to days 20-24 of the menstrual cycle, aligned by cell types.

**Supplementary File 6**. Receptivity-associated genes and HRT genes, differentially expressed in AMA samples. **a.** Boxplots of AMA significant genes associated with WOI in RNA-seq based publications. **b**. UMAP of YMA and AMA samples based on WOI genes. **c.** Boxplots of AMA significant genes associated with HRT. **b**. UMAP of YMA and AMA samples based on HRT genes.

**Supplementary Figures 1-5.** Significant molecular interaction networks predicted by IPA.

## References

1. Cornelis Lambalk. Editor’s Choice: Delayed childbearing and medically assisted reproduction. Human Reproduction. Published online 2022.

2. Moore L, Leongamornlert D, Coorens THH, et al. The mutational landscape of normal human endometrial epithelium. Nature. 2020;580(7805):640-646.

3. Abdalla HI, Wren ME, Thomas A, Korea L. Age of the uterus does not affect pregnancy or implantation rates; a study of egg donation in women of different ages sharing oocytes from the same donor. Human Reproduction. 1997;12(4):827–829.

4. Flamigni C, Borini A, Violini F, Bianchi L, Serrao L. Infertility: Oocyte donation: comparison between recipients from different age groups. Human Reproduction. 1993;8(12):2088–2092.

5. Soares SR, Troncoso C, Bosch E, et al. Age and Uterine Receptiveness: Predicting the Outcome of Oocyte Donation Cycles. The Journal of Clinical Endocrinology & Metabolism. 2005;90(7):4399–4404.

6. Perkins KM, Boulet SL, Jamieson DJ, Kissin DM. Trends and outcomes of gestational surrogacy in the United States. Fertility and Sterility. 2016;106(2):435–442.e2.

7. Devesa-Peiro A, Sebastian-Leon P, Parraga-Leo A, Pellicer A, Diaz-Gimeno P. Breaking the ageing paradigm in endometrium: endometrial gene expression related to cilia and ageing hallmarks in women over 35 years. Human Reproduction. 2022;37(4):762–776.

8. Suhorutshenko M, Kukushkina V, Velthut-Meikas A, et al. Endometrial receptivity revisited: endometrial transcriptome adjusted for tissue cellular heterogeneity. Human Reproduction. 2018;33(11):2074–2086.

9. Hu S, Yao G, Wang Y, et al. Transcriptomic Changes During the Pre-Receptive to Receptive Transition in Human Endometrium Detected by RNA-Seq. The Journal of Clinical Endocrinology & Metabolism. 2014;99(12):E2744–E2753.

10. Sigurgeirsson B, Åmark H, Jemt A, et al. Comprehensive RNA sequencing of healthy human endometrium at two time points of the menstrual cycle. Biol Reprod. 2017;96(1):24–33.

11. Altmäe S, Tamm-Rosenstein K, Esteban FJ, et al. Endometrial transcriptome analysis indicates superiority of natural over artificial cycles in recurrent implantation failure patients undergoing frozen embryo transfer. Reproductive BioMedicine Online. 2016;32(6):597–613.

12. Niu G, Hao J, Sheng S, Wen F. Role of TLbox genes in cancer, epithelialLmesenchymal transition, and cancer stem cells. J of Cellular Biochemistry. 2022;123(2):215–230.

13. Arribas J, Cajuso T, Rodio A, Marcos R, Leonardi A, Velázquez A. NF-κB Mediates the Expression of TBX15 in Cancer Cells. Castresana JS, ed. PLoS ONE. 2016;11(6):e0157761.

14. Owusu-Akyaw A, Krishnamoorthy K, Goldsmith LT, Morelli SS. The role of mesenchymal– epithelial transition in endometrial function. Human Reproduction Update. 2019;25(1):114–133.

15. Li H, Cao G, Zhang N, et al. RBP4 regulates trophoblastic cell proliferation and invasion via the PI3K/AKT signaling pathway. Mol Med Report. Published online July 3, 2018.

16. Pavone ME, Malpani S, Dyson M, Bulun SE. Altered retinoid signaling compromises decidualization in human endometriotic stromal cells. Reproduction. 2017;154(3):207–216.

17. Aghajanova L, Altmae S, Stavreus-Evers A, Giudice LC. Stanniocalcin-1 in Human Endometrium. Fertility and Sterility. 2015;103(2):e6–e7.

18. Aghajanova L, Altmäe S, Kasvandik S, Salumets A, Stavreus-Evers A, Giudice LC. Stanniocalcin-1 expression in normal human endometrium and dysregulation in endometriosis. Fertility and Sterility. 2016;106(3):681–691.e1.

19. for the ADVANCE Collaborative group, Zoungas S, Woodward M, et al. Impact of age, age at diagnosis and duration of diabetes on the risk of macrovascular and microvascular complications and death in type 2 diabetes. Diabetologia. 2014;57(12):2465–2474.

20. Haouzi D, Mahmoud K, Fourar M, et al. Identification of new biomarkers of human endometrial receptivity in the natural cycle. Human Reproduction. 2008;24(1):198–205.

21. Turkyilmaz E, Guner H, Erdem M, et al. NLF2 gene expression in the endometrium of patients with implantation failure after IVF treatment. Gene. 2012;508(1):140–143.

22. Martin N, Beach D, Gil J. Ageing as developmental decay: insights from p16INK4a. Trends in Molecular Medicine. 2014;20(12):667–674.

23. Parvanov D, Ganeva R, Vidolova N, Stamenov G. Decreased number of p16-positive senescent cells in human endometrium as a marker of miscarriage. J Assist Reprod Genet. 2021;38(8):2087–2095.

24. Narita M, Nuñez S, Heard E, et al. Rb-Mediated Heterochromatin Formation and Silencing of E2F Target Genes during Cellular Senescence. Cell. 2003;113(6):703–716.

25. Teissier T, Boulanger E, Cox LS. Interconnections between inflammageing and immunosenescence during ageing. Cells. 2022;11(3):359.

26. Brighton PJ, Maruyama Y, Fishwick K, et al. Clearance of senescent decidual cells by uterine natural killer cells in cycling human endometrium. eLife. 2017;6:e31274.

27. Lujambio A. To clear, or not to clear (senescent cells)? That is the question: Clearance of senescent cells. BioEssays. 2016;38:S56–S64.

28. Baker DJ, Childs BG, Durik M, et al. Naturally occurring p16Ink4a-positive cells shorten healthy lifespan. Nature. 2016;530(7589):184-189.

29. Kagawa S, Natsuizaka M, Whelan KA, et al. Cellular senescence checkpoint function determines differential Notch1-dependent oncogenic and tumor-suppressor activities. Oncogene. 2015;34(18):2347–2359.

30. Tasca A, Helmstädter M, Brislinger MM, Haas M, Mitchell B, Walentek P. Notch signaling induces either apoptosis or cell fate change in multiciliated cells during mucociliary tissue remodeling. Developmental Cell. 2021;56(4):525–539.e6.

31. Vassilieva IO, Reshetnikova GF, Shatrova AN, et al. Senescence-messaging secretome factors trigger premature senescence in human endometrium-derived stem cells. Biochemical and Biophysical Research Communications. 2018;496(4):1162–1168.

32. Singer D, Thamm K, Zhuang H, et al. PromininL1 controls stem cell activation by orchestrating ciliary dynamics. EMBO J. 2019;38(2).

33. Horree N, Heintz APM, Sie-Go DMDS, van Diest PJ. p16 is Consistently Expressed in Endometrial Tubal Metaplasia. Analytical Cellular Pathology. 2007;29(1):37–45.

34. Klaes R, Friedrich T, Spitkovsky D, et al. Overexpression of p16INK4A as a specific marker for dysplastic and neoplastic epithelial cells of the cervix uteri. Int J Cancer. 2001;92(2):276–284.

35. Haider S, Gamperl M, Burkard TR, et al. Estrogen Signaling Drives Ciliogenesis in Human Endometrial Organoids. Endocrinology. 2019;160(10):2282–2297.

36. Serra CFH, Liu H, Qian J, Mori M, Lu J, Cardoso WV. Prominin 1 and Notch regulate ciliary length and dynamics in multiciliated cells of the airway epithelium. iScience. 2022;25(8):104751.

37. More IAR, Masterton RG. The role of oestrogen in the control of ciliated cells of the human endometrium. Reproduction. 1976;47(1):19–24.

38. Masterton R, Armstrong EM, More IAR. The cyclical variation in the percentage of ciliated cells in the human endometrium. Reproduction. 1975;42(3):537–540.

39. Hale GE, Zhao X, Hughes CL, Burger HG, Robertson DM, Fraser IS. Endocrine Features of Menstrual Cycles in Middle and Late Reproductive Age and the Menopausal Transition Classified According to the Staging of Reproductive Aging Workshop (STRAW) Staging System. The Journal of Clinical Endocrinology & Metabolism. 2007;92(8):3060–3067.

40. Li MQ, Yao MN, Yan JQ, et al. The decline of pregnancy rate and abnormal uterine responsiveness of steroid hormones in aging mice. Reproductive Biology. 2017;17(4):305–311.

41. Wang W, Vilella F, Alama P, et al. Single-cell transcriptomic atlas of the human endometrium during the menstrual cycle. Nat Med. 2020;26(10):1644–1653.

42. Patir A, Fraser AM, Barnett MW, et al. The transcriptional signature associated with human motile cilia. Sci Rep. 2020;10(1):10814.

43. Ide T, Twan WK, Lu H, et al. CFAP53 regulates mammalian cilia-type motility patterns through differential localization and recruitment of axonemal dynein components. Dutcher SK, ed. PLoS Genet. 2020;16(12):e1009232.

44. Mijnes J, Bringezu S, Berger J, et al. SPAG6 Promotes Cell Migration and Induces Epithelial-to-Mesenchymal Transition in Luminal Breast Cancer Cells. Cell Biology; 2022.

45. Ho JC, Chan KN, Hu WH, et al. The Effect of Aging on Nasal Mucociliary Clearance, Beat Frequency, and Ultrastructure of Respiratory Cilia. Am J Respir Crit Care Med. 2001;163(4):983–988.

46. Díaz-Gimeno P, Horcajadas JA, Martínez-Conejero JA, et al. A genomic diagnostic tool for human endometrial receptivity based on the transcriptomic signature. Fertility and Sterility. 2011;95(1):50–60.e15.

47. Altmäe S, Koel M, Võsa U, et al. Meta-signature of human endometrial receptivity: a meta-analysis and validation study of transcriptomic biomarkers. Sci Rep. 2017;7(1):10077.

48. Enciso M, Carrascosa JP, Sarasa J, et al. Development of a new comprehensive and reliable endometrial receptivity map (ER Map/ER Grade) based on RT-qPCR gene expression analysis. Human Reproduction. 2018;33(2):220–228.

49. Haouzi D. Endometrial receptivity under hormone replacement therapy in oocyte-donation recipient patients: transcriptomic approach. MRAJ. 2015;2(1).

50. Zeng H, Fu Y, Shen L, Quan S. Integrated Analysis of Multiple Microarrays Based on Raw Data Identified Novel Gene Signatures in Recurrent Implantation Failure. Front Endocrinol. 2022;13:785462.

51. Díaz-Gimeno P, Ruiz-Alonso M, Sebastian-Leon P, Pellicer A, Valbuena D, Simón C. Window of implantation transcriptomic stratification reveals different endometrial subsignatures associated with live birth and biochemical pregnancy. Fertility and Sterility. 2017;108(4):703–710.e3.

52. Shen TL, Han DC, Guan JL. Association of Grb7 with Phosphoinositides and Its Role in the Regulation of Cell Migration. Journal of Biological Chemistry. 2002;277(32):29069–29077.

53. Wirstlein PK, Mikołajczyk M, Skrzypczak J. Correlation of the expression of heparanase and heparin-binding EGF-like growth factor in the implantation window of nonconceptual cycle endometrium. Folia Histochem Cytobiol. 2013;51(2):127–134.

54. D’Souza SS, Daikoku T, Farach-Carson MC, Carson DD. Heparanase Expression and Function During Early Pregnancy in Mice1. Biology of Reproduction. 2007;77(3):433–441.

55. Revel A, Helman A, Koler M, et al. Heparanase improves mouse embryo implantation. Fertility and Sterility. 2005;83(3):580–586.

56. Meltsov A, Saare M, Teder H, et al. Targeted Gene Expression Profiling for Accurate Endometrial Receptivity Testing. Genetic and Genomic Medicine; 2022.

57. Wu H, Wang C, Wu Z. PROPER: comprehensive power evaluation for differential expression using RNA-seq. Bioinformatics. 2015;31(2):233–241.

58. Livak KJ, Schmittgen TD. Analysis of Relative Gene Expression Data Using Real-Time Quantitative PCR and the 2−ΔΔCT Method. Methods. 2001;25(4):402–408.

59. Fischer AH, Jacobson KA, Rose J, Zeller R. Hematoxylin and Eosin Staining of Tissue and Cell Sections. Cold Spring Harb Protoc. 2008;2008(5):pdb.prot4986.

60. Noyes RW, Hertig AT, Rock J. Dating the Endometrial Biopsy. Fertility and Sterility. 1950;1(1):3–25.

61. Schindelin J, Arganda-Carreras I, Frise E, et al. Fiji: an open-source platform for biological-image analysis. Nat Methods. 2012;9(7):676–682.

62. Fuhrich DG, Lessey BA, Savaris RF. Comparison of HSCORE assessment of endometrial beta3 integrin subunit expression with digital HSCORE using computerized image analysis (ImageJ). Anal Quant Cytopathol Histpathol. 2013;35(4):210–216.

63. Tsoucas D, Dong R, Chen H, Zhu Q, Guo G, Yuan GC. Accurate estimation of cell-type composition from gene expression data. Nat Commun. 2019;10(1):2975.

