## Supplementary Figures for "Aging affects ciliated cells development in the human endometrial epithelium"

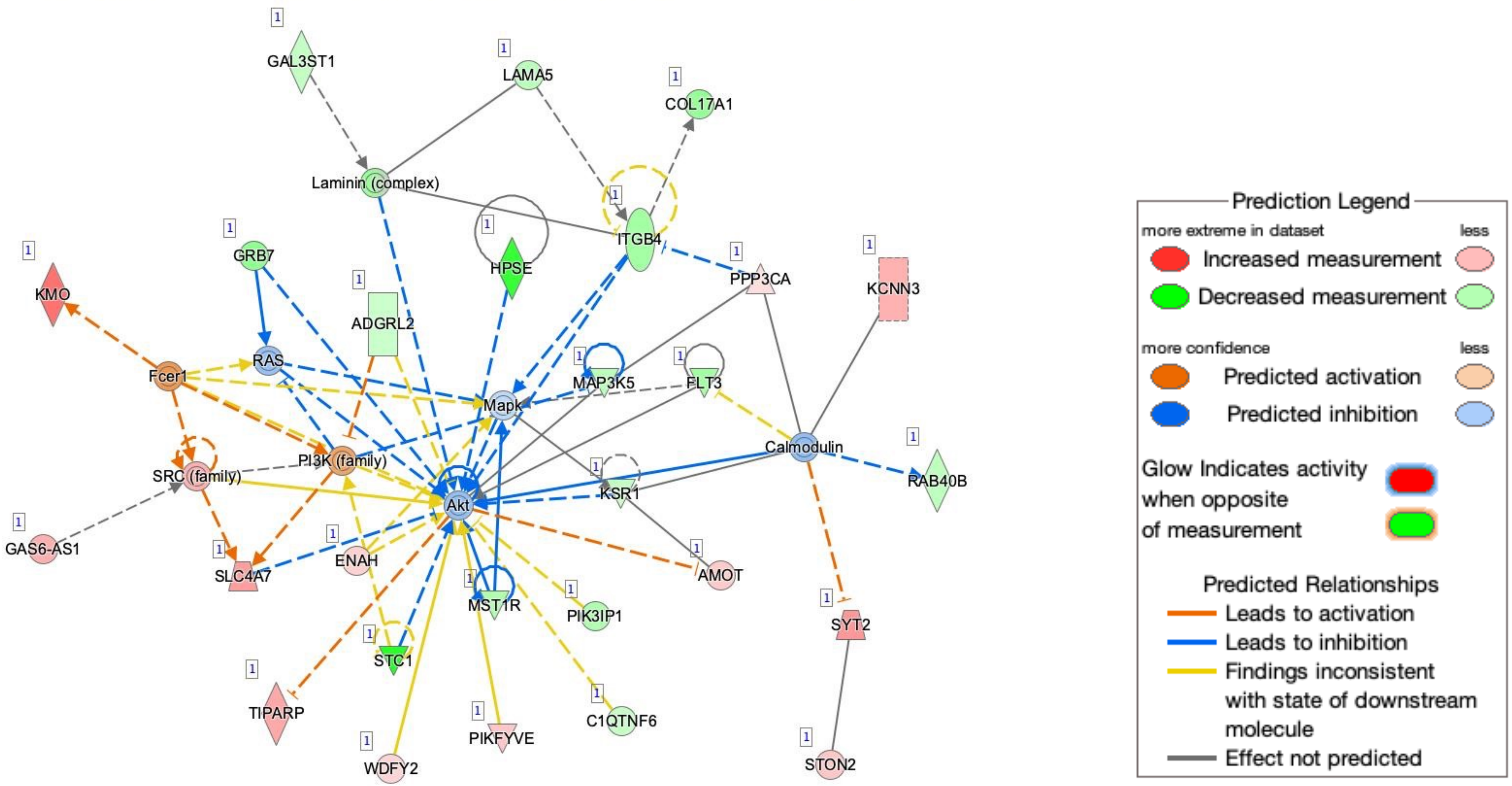


**Supplementary Fig.1.** IPA molecular interaction network 1: Cellular Movement

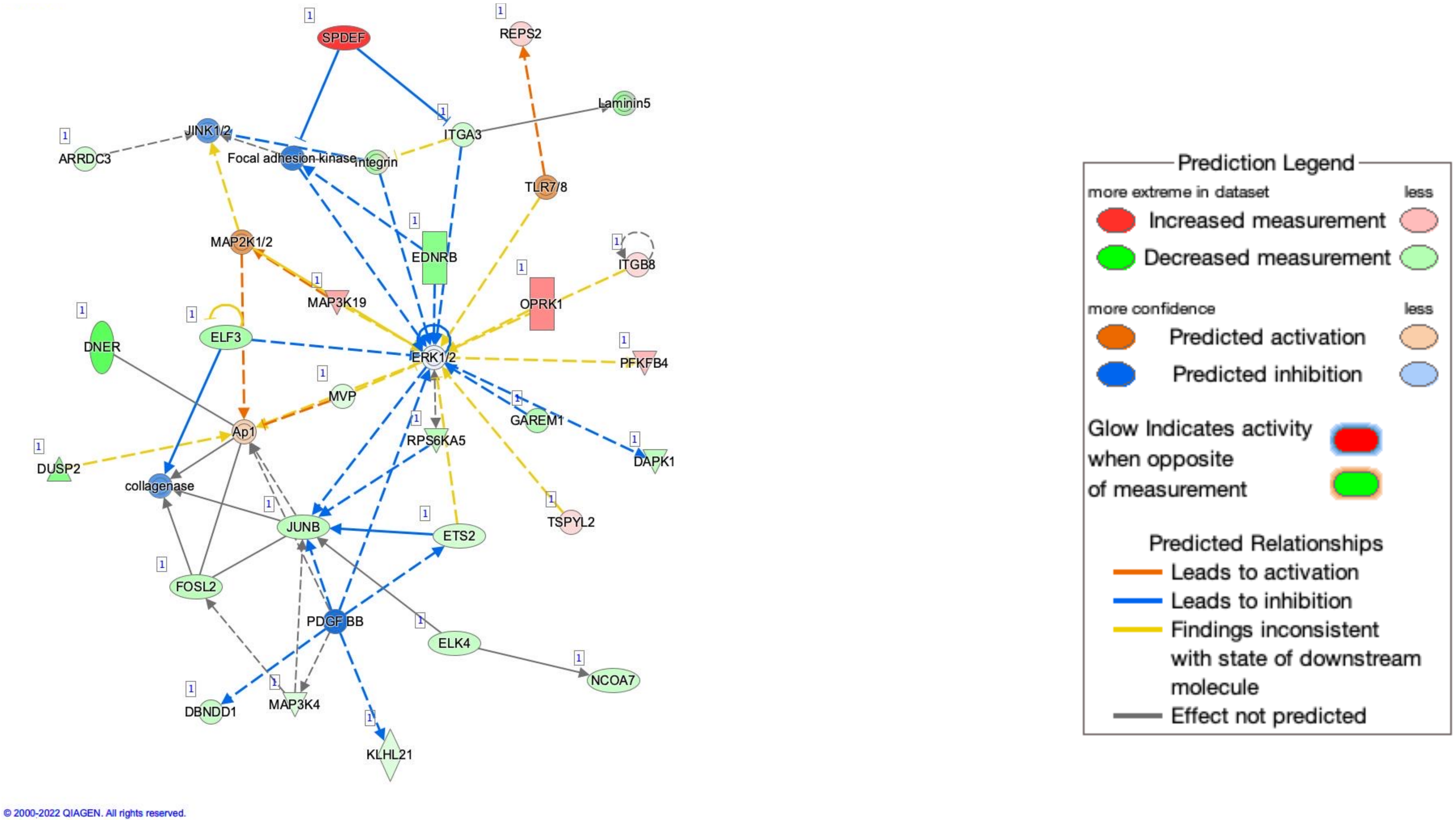


**Supplementary Fig.2.** IPA molecular interaction network 2. Cellular Development

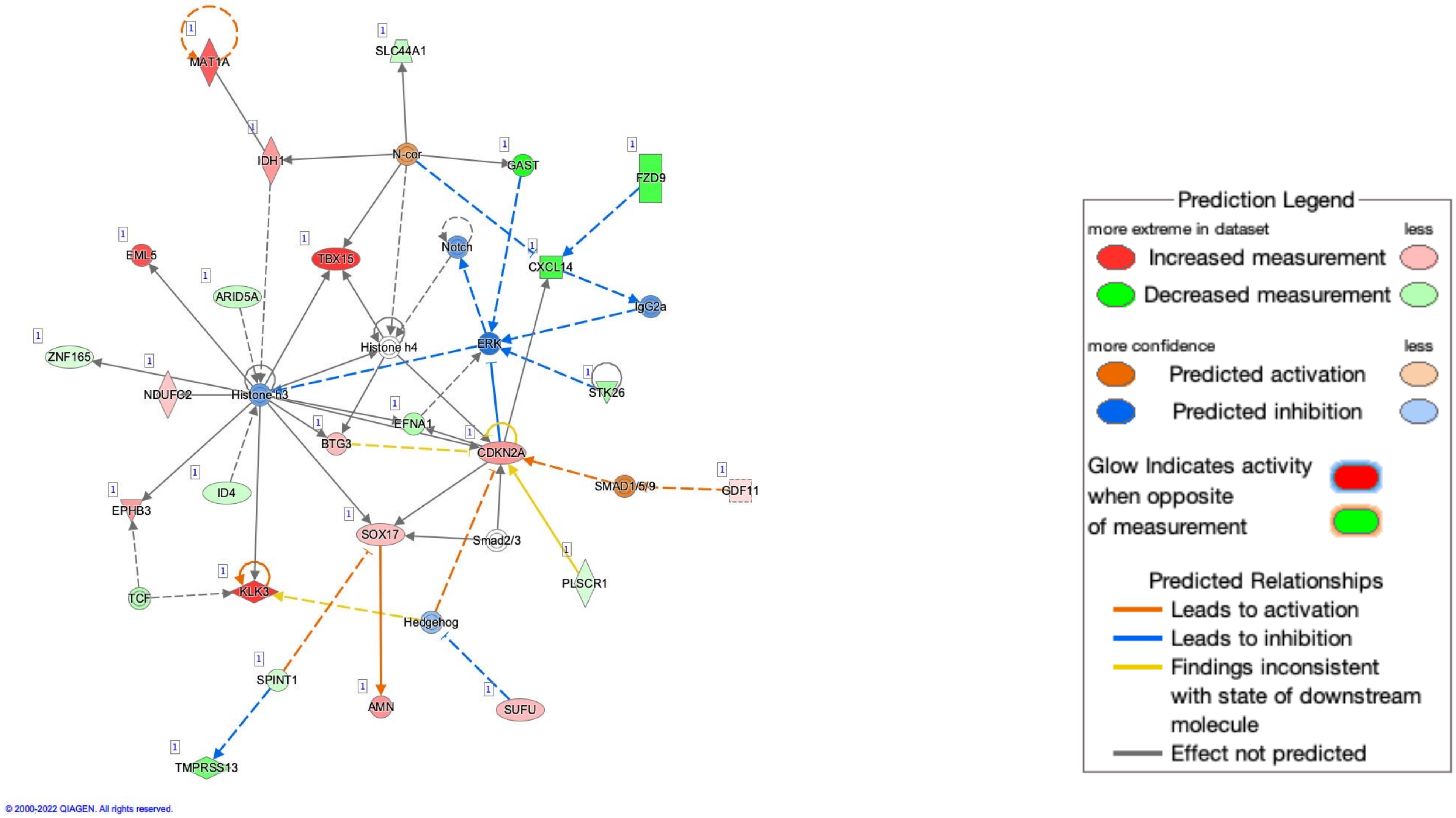


**Supplementary Fig.3.** IPA Network 3. Cancer

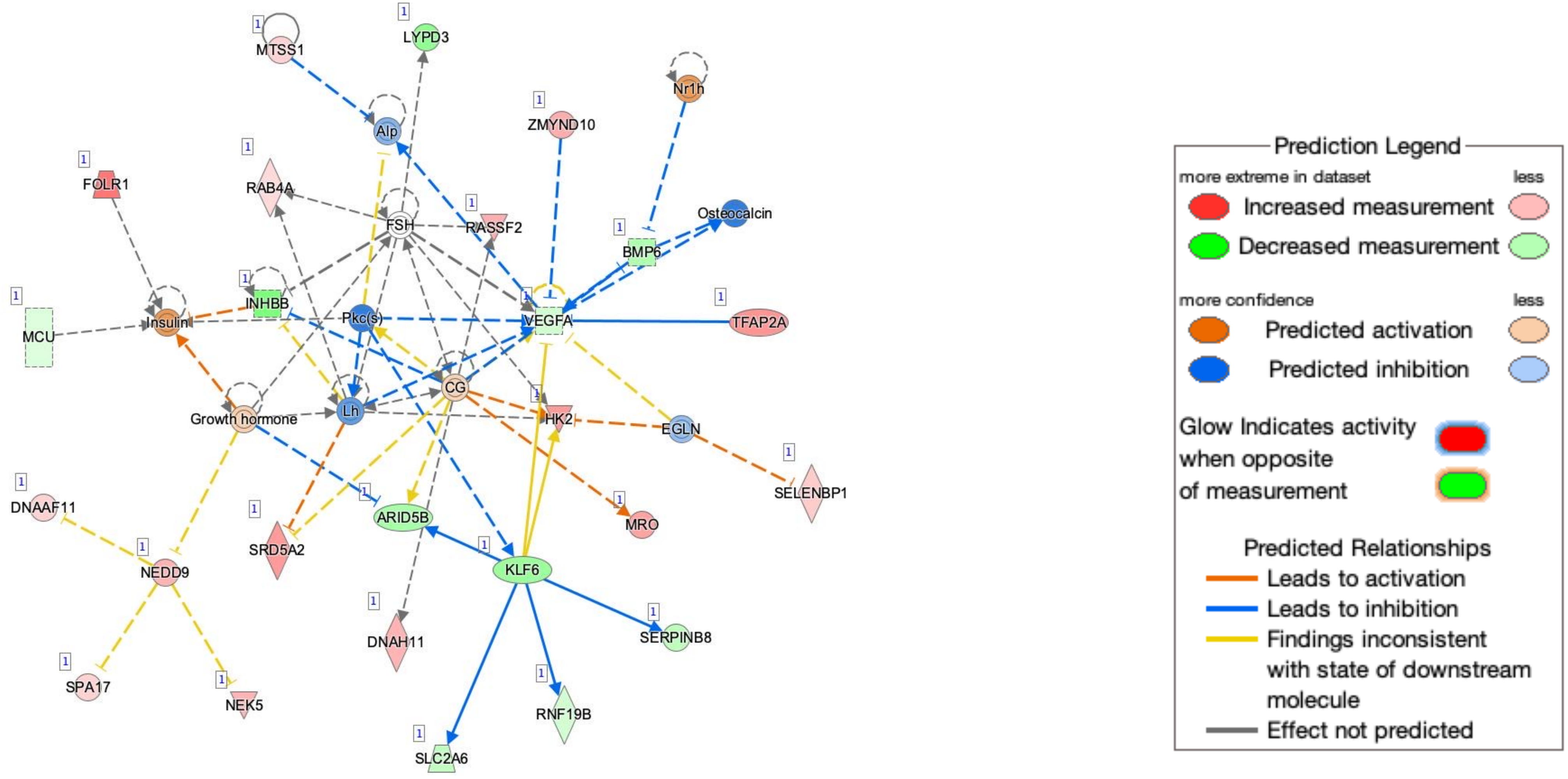


**Supplementary Fig.4.** IPA Network 4. Cardiovascular Disease

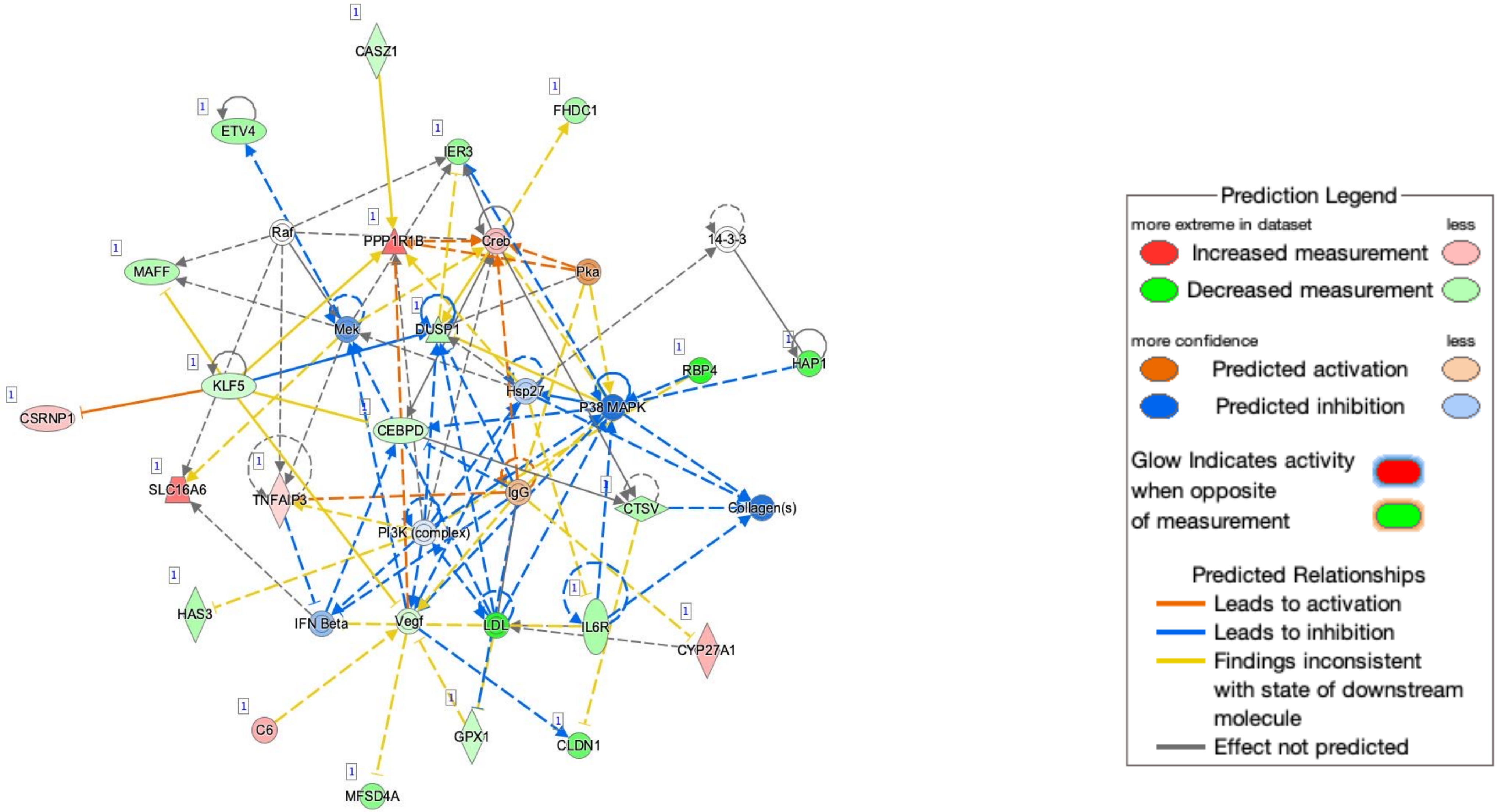


© 2000-2022 QIAGEN. All rights reserved.

**Supplementary Fig.5.** IPA Network 5. Cardiac Enlargement, Cardiovascular Disease
