## Supplementary File 1 for "Aging affects ciliated cells development in the human endometrial epithelium"

| Sample | Clean reads (after QC) | GC (%) | Aligned reads (%) | Total aligned reads (million) |
|---|---|---|---|---|
| RNA108 | 34M | 47.0 | 91.94 | 31M |
| RNA138 | 37M | 48.0 | 92.37 | 34M |
| RNA145 | 33M | 47.0 | 90.24 | 29M |
| RNA148 | 44M | 48.0 | 91.45 | 40M |
| RNA164 | 42M | 48.0 | 90.82 | 38M |
| RNA166 | 32M | 48.0 | 91.85 | 30M |
| RNA191 | 35M | 48.0 | 92.01 | 32M |
| RNA208 | 40M | 48.0 | 91.69 | 36M |
| RNA268 | 37M | 48.0 | 91.92 | 34M |
| RNA358 | 43M | 48.0 | 91.12 | 40M |
| RNA382 | 31M | 49.0 | 91.42 | 28M |
| RNA473 | 32M | 48.0 | 92.56 | 29M |
| RNA516 | 36M | 48.0 | 92.13 | 33M |
| RNA562 | 32M | 49.0 | 92.12 | 29M |
| RNA638 | 33M | 48.0 | 91.63 | 31M |
| RNA70 | 35M | 48.0 | 91.14 | 32M |
| RNA705 | 40M | 48.0 | 91.38 | 36M |
| RNA709 | 33M | 48.0 | 91.75 | 30M |
| RNA83 | 32M | 47.0 | 91.59 | 29M |
| RNA859 | 36M | 49.0 | 91.93 | 33M |
| RNA868 | 62M | 49.0 | 91.51 | 57M |
| RNA886 | 43M | 49.0 | 91.54 | 39M |
| RNA893 | 37M | 48.0 | 92.29 | 34M |
| RNA989 | 27M | 48.0 | 90.94 | 24M |

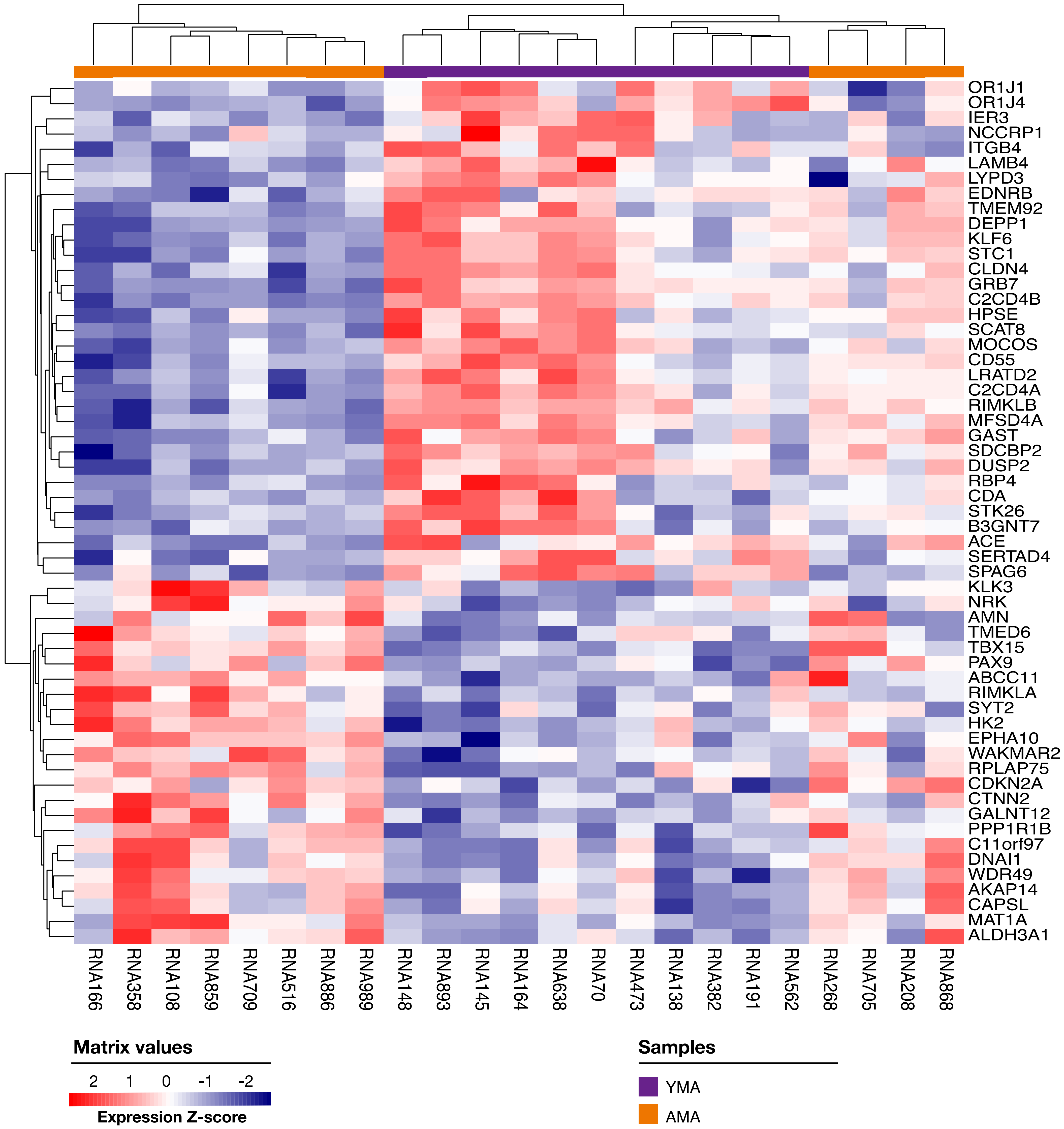
OR1J1
OR1J4
IER3
NCCRP1
ITGB4
LAMB4
LYPD3
EDNRB
TMEM92
DEPP1
KLF6
STC1
CLDN4
GRB7
C2CD4B
HPSE
SCAT8
MOCOS
CD55
LRATD2
C2CD4A
RIMKLB
MFSD4A
GAST
SDCBP2
DUSP2
RBP4
CDA
STK26
B3GNT7
ACE
SERTAD4
SPAG6
KLK3
NRK
AMN
TMED6
TBX15
PAX9
ABCC11
RIMKLA
SYT2
HK2
EPHA10
WAKMAR2
RPLAP75
CDKN2A
CTNN2
GALNT12
PPP1R1B
C11orf97
DNAI1
WDR49
AKAP14
CAPSL
MAT1A
ALDH3A1
RNA166
RNA358
RNA108
RNA859
RNA709
RNA516
RNA886
RNA989
RNA148
RNA893
RNA145
RNA164
RNA638
RNA70
RNA473
RNA138
RNA382
RNA191
RNA562
RNA268
RNA705
RNA208
RNA868
Matrix values
2
1
0
-1
-2
Expression Z-score
Samples
YMA
AMA

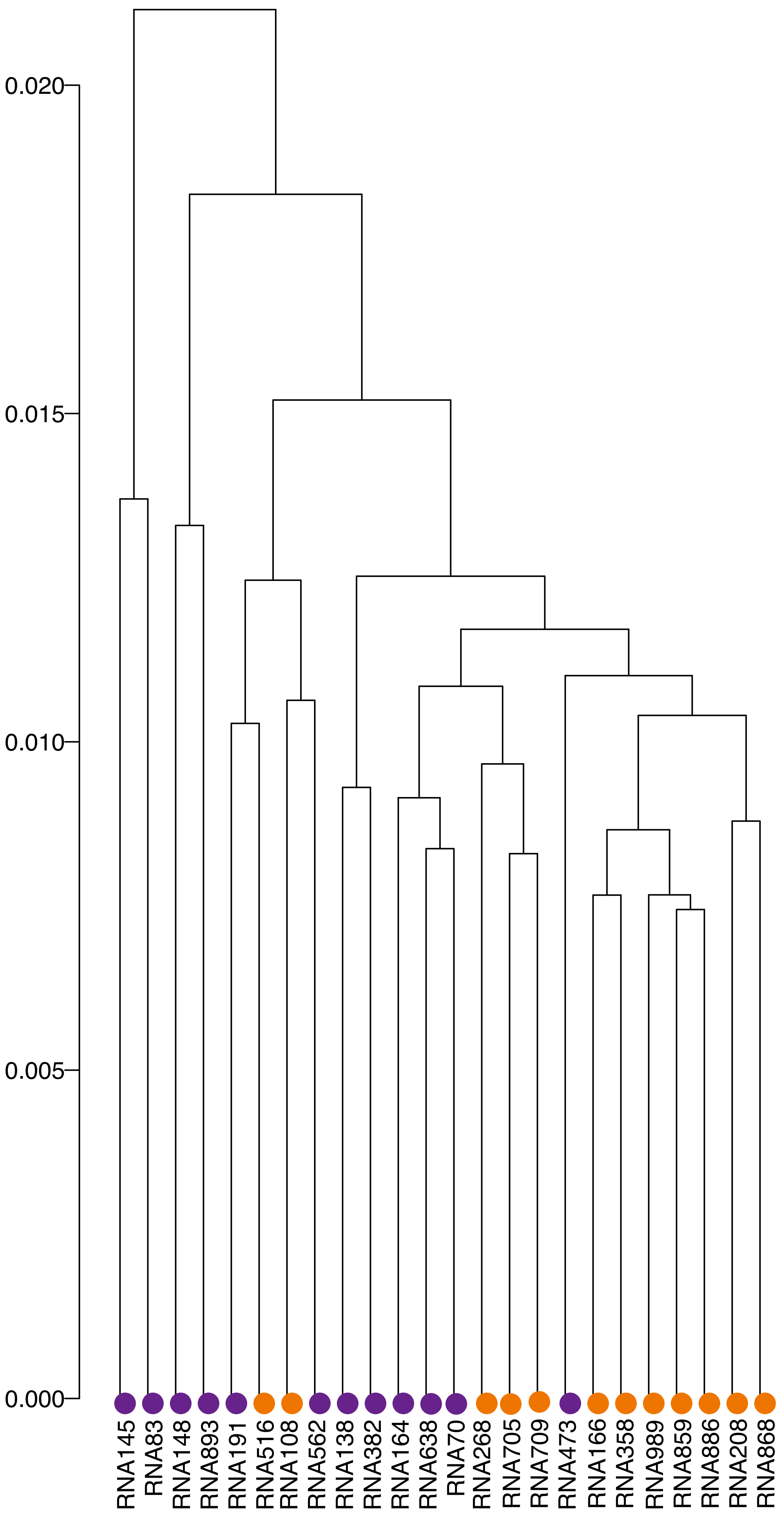
0.020
0.015
0.010
0.005
0.000
RNA145
RNA83
RNA148
RNA893
RNA191
RNA516
RNA108
RNA562
RNA138
RNA382
RNA164
RNA638
RNA70
RNA268
RNA705
RNA709
RNA473
RNA166
RNA358
RNA989
RNA859
RNA886
RNA208
RNA868
