## Supplementary figures and images for "Aging affects ciliated cells development in the human endometrial epithelium"

### Supplemenatary File 5

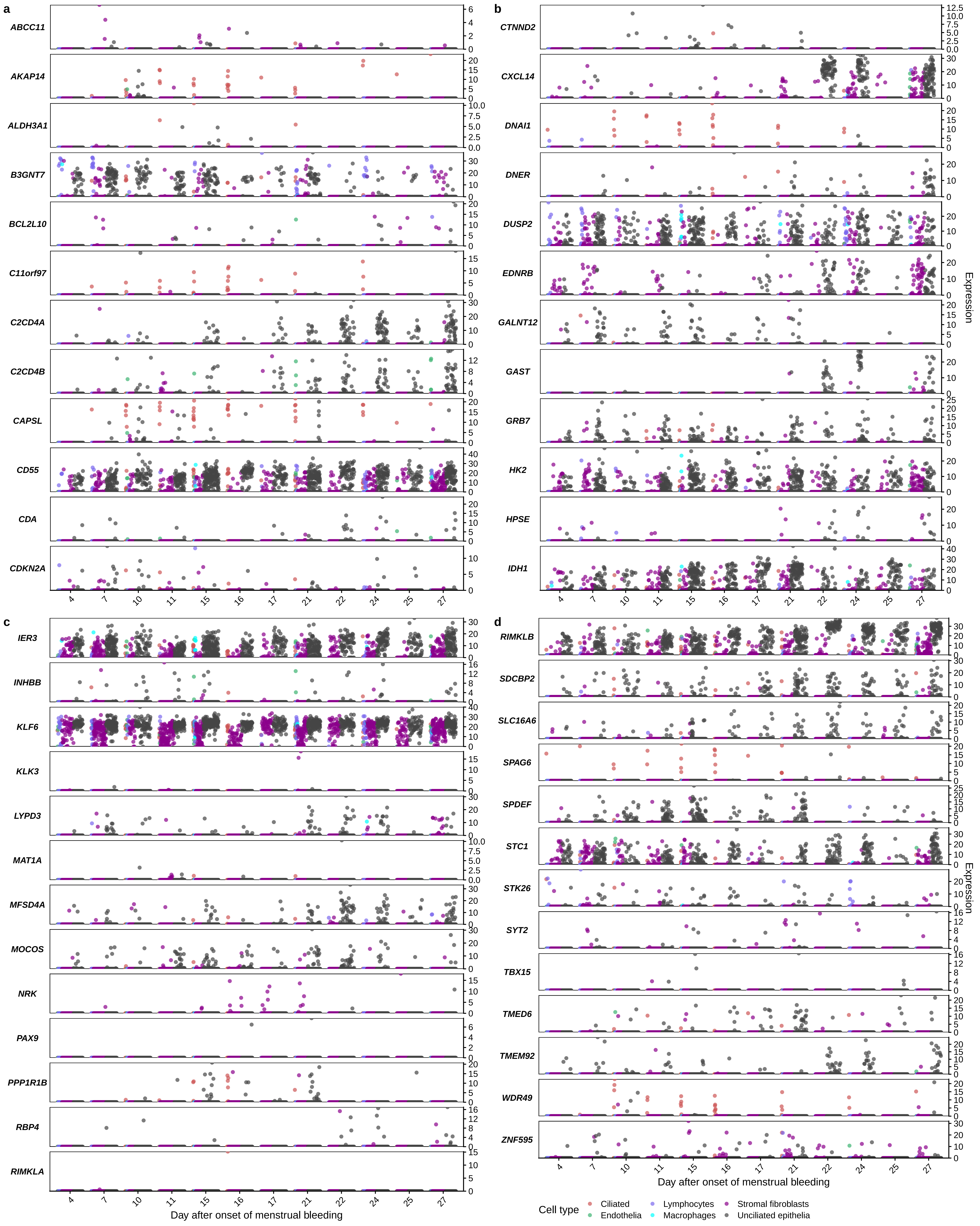

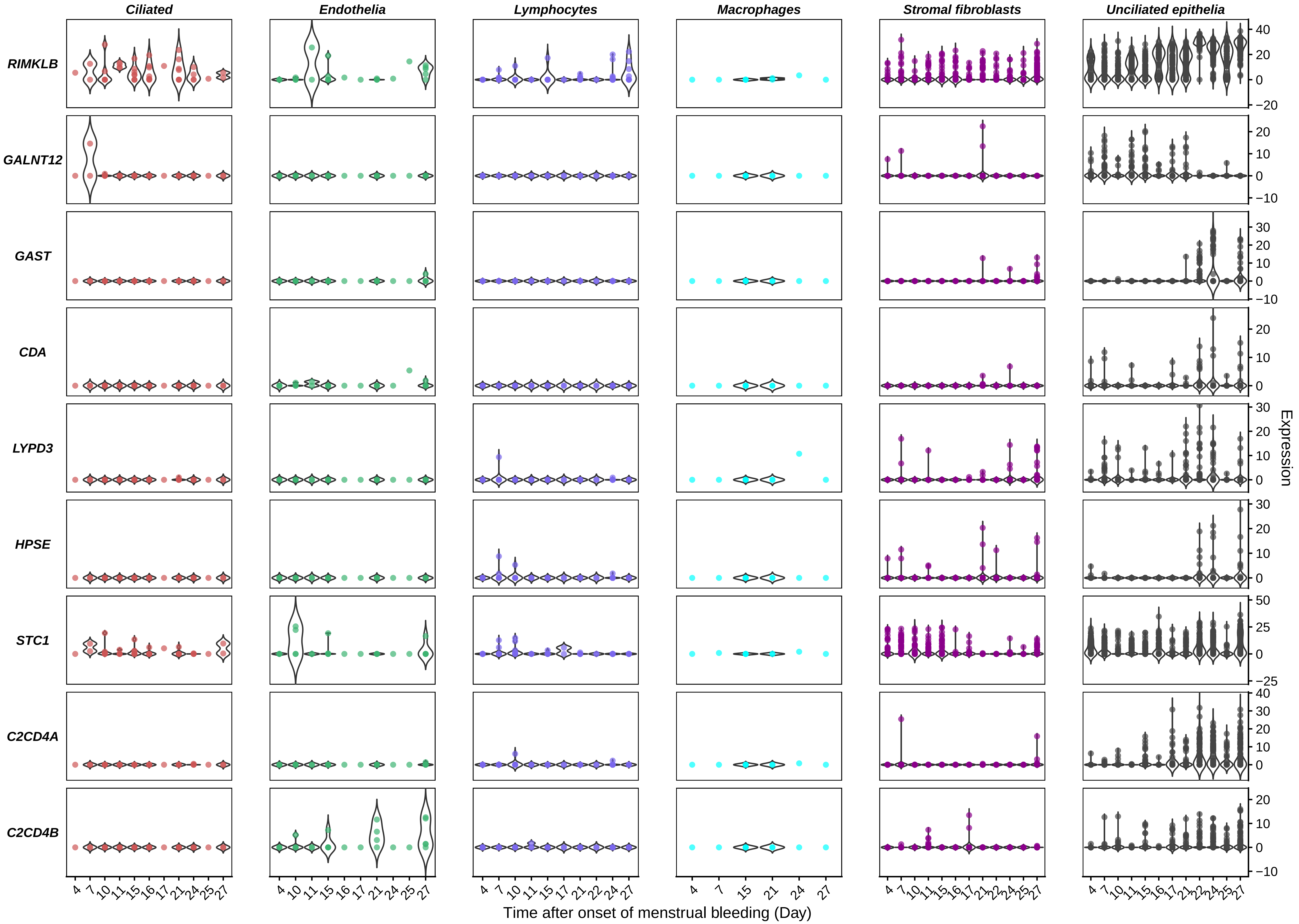

### Supplementary File 6

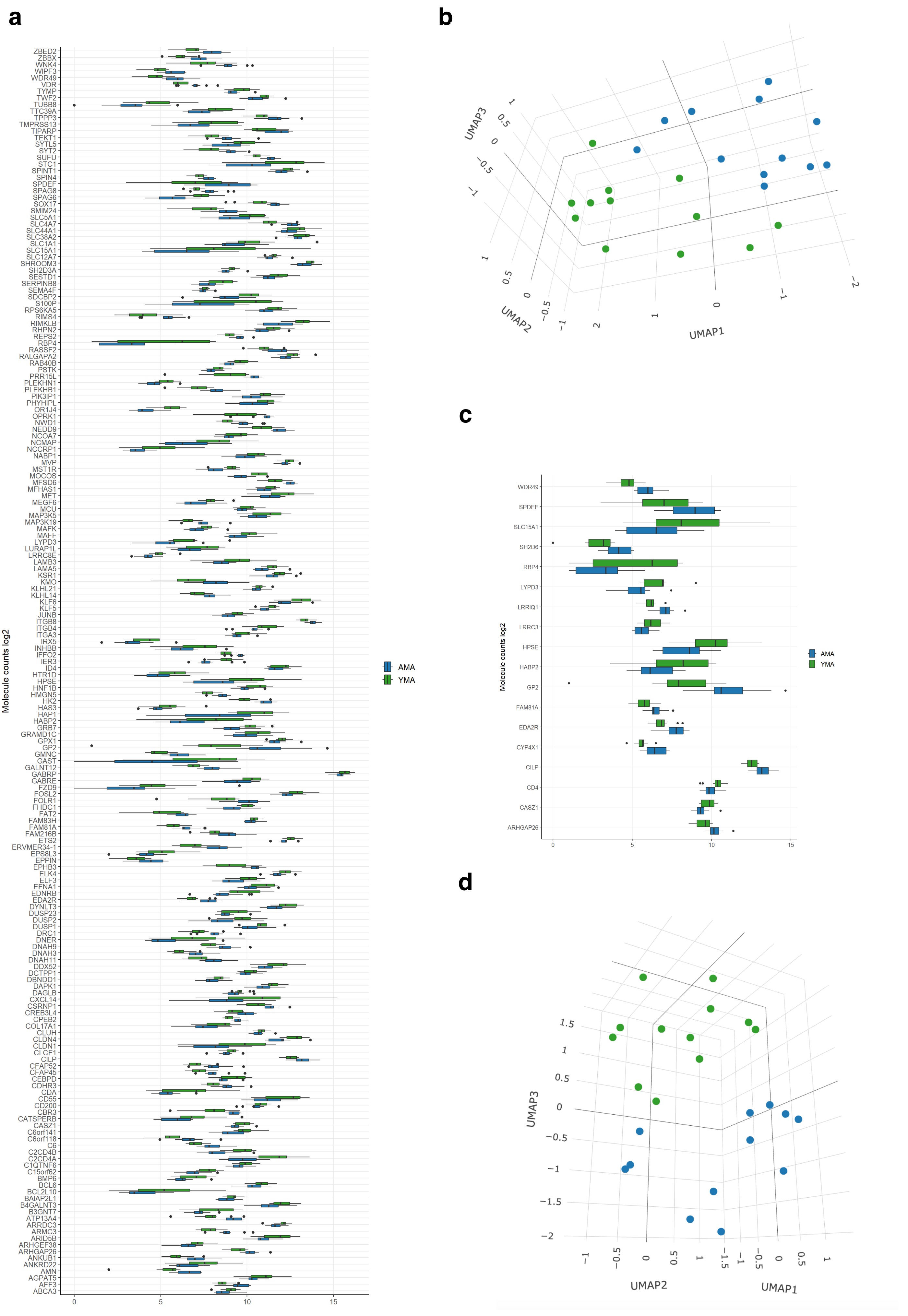
